# JointMR: A joint likelihood-based approach for causal effect estimation in overlapping Mendelian Randomization studies

**DOI:** 10.64898/2025.12.18.25342634

**Authors:** Sijia Wu, Lei Hou, Zhongshang Yuan, Xiaoru Sun, Yuanyuan Yu, Hao Chen, Lin Huang, Fuzhong Xue, Hongkai Li

## Abstract

The integration of causal effect estimates from multiple Mendelian Randomization studies has become increasingly popular. However, the presence of overlapping databases compromises traditional meta-analysis, leading to inflated variance and reduced statistical power. Here, we propose JointMR, a joint likelihood-based approach designed to integrate multiple GWAS summary databases while explicitly accounting for the covariance matrix of the Wald ratio estimates. Specifically, to accommodate potential cross-study heterogeneity, JointMR incorporates both fixed-effect and random-effects models. Simulations demonstrated that JointMR provides unbiased estimates with higher statistical power and superior Type I error control compared to conventional meta-analysis methods of standard MR estimates (e.g., IVW), especially as database correlation increases. In a real-data application examining total cholesterol, HDL-C, LDL-C and triglycerides on type 2 diabetes, JointMR resolved contradictions seen in standard approaches, generating stable and biologically plausible estimates. In conclusion, JointMR overcomes critical limitations of existing methods, offering a more powerful and reliable tool for robust causal inference from the growing repository of GWAS summary statistics.

## Introduction

Mendelian randomization (MR) has established itself as a cornerstone for causal inference in observational epidemiology, leveraging genetic variants as instrumental variables (IVs) to mitigate confounding and reverse causation [1–3]. The proliferation of genome-wide association study (GWAS) summary statistics, such as the UK Biobank (UKB), international consortia [4, 5], FinnGen [6] and GWAS catalog [7], has further propelled the application of MR, enabling the investigation of putative causal relationships across a vast array of complex traits and diseases. Concurrently, meta-analysis stands as a fundamental statistical paradigm for enhancing power and precision by combining results from multiple independent studies [8]. However, its application in MR synthesis is driven by a more urgent need: different MR studies, often using distinct databases or methodologies, frequently report inconsistent or even conflicting estimates for the same causal question. For instance, given the heterogeneous design of existing MR studies on Osteoarthritis (OA), reported causal estimates remain inconsistent, obscuring the true biological effects [9–11]. A formal meta-analysis is therefore crucial to synthesize this heterogeneous evidence, resolve discrepancies, and derive a single, robust causal inference.

The methodology for synthesizing MR studies, however, has not kept pace with the evolution of MR itself (e.g., Inverse-Variance Weighted (IVW) [12], MR-Egger [13], Weighted Median (WME) [14]). In practice, researchers have adopted two main simplified approaches. For instance, in large-scale consortium efforts, such as the prominent analysis of Lp(a) and coronary heart disease [15], the common strategy is to first combine exposure or outcome GWAS using traditional tools (like METAL [16]) to maximize statistical power before MR is performed. Alternatively, in systematic reviews aiming to synthesize all available evidence—for instance, the comprehensive review by Ho et al. [17] on osteoarthritis risk factors—the approach is to pool the final causal estimates from multiple, seemingly independent MR studies [18]. These common practices can be categorized as “upstream” meta-analysis and “downstream” meta-analysis, respectively.

Neither strategy is statistically optimal. The “downstream” approach of pooling final estimates, in particular, faces two major challenges. First, it is highly susceptible to publication bias; systematic reviews can only synthesize published MR studies, which are often skewed towards statistically significant findings, while true null effects, though equally valid, remain unpublished [19–20]. This selective reporting can lead to a distorted and overestimated causal inference. Second, as a practical problem in many published MR meta-analyses, this strategy fails to leverage the full data structure and, critically, ignores the complex dependency (i.e., correlation) inherent in the design. For instance, in large-scale umbrella reviews, such as the one by Kim et al. [21] on adiposity and cardiovascular outcomes and the meta-analysis by Chen et al. [22] on telomere length, many of the ‘independent’ MR studies being synthesized actually drew their outcome data from the same large consortium (e.g., UK Biobank). This sample overlap induces a non-zero covariance between their MR estimates. Ignoring this covariance, as standard meta-analysis models [8] do, violates core statistical assumptions and leads to biased standard errors, suboptimal efficiency, and potentially invalidated statistical inference.

The limitations of these two simplified strategies jointly point to a clear research gap: the lack of a unified meta-analysis framework specifically designed for the *N* exposure and *M* outcome setting. The primary challenge lies in modeling the complex, block-structured covariance matrix that arises when estimates share a common outcome GWAS database. Traditional meta-analysis models are ill-equipped to handle this systemic dependency, as they typically assume a simple diagonal covariance matrix (i.e., independence between all MR estimates). This assumption is clearly violated in the multiple databases scenario and inevitably leads to biased standard errors and suboptimal statistical efficiency. Furthermore, this challenge is compounded by the pervasive issue of horizontal pleiotropy. While existing robust MR methods can handle pleiotropy within a single exposure-outcome pair, they were not designed for this multi-database framework [23–24]. A key technical hurdle, therefore, is to robustly address horizontal pleiotropy across these multiple databases while simultaneously accounting for the aforementioned block-structured covariance. An integrated method capable of addressing both core issues is currently lacking.

Here, we introduce a novel statistical approach named JointMR that directly addresses this gap. Our method is designed to integrate summary-level data from *N* independent GWAS databases for an exposure and *M* independent databases for an outcome within a single, unified likelihood-based model. The core innovation lies in explicitly modeling the covariance matrix as a structured block matrix to account for correlations between estimates sharing the same outcome database. We develop both fixed-effect and random-effects models, employing maximum likelihood estimation for inference. To ensure robustness against horizontal pleiotropy, we incorporate an adjustment using MR-Egger estimates, deriving a modified Wald ratio that corrects for pleiotropic effects before meta-analysis. By simultaneously harnessing all available data and correctly accounting for its complex structure, our approach provides a more powerful and statistically rigorous approach for causal inference, effectively bridging the gap between advanced MR methodology and the realities of multi-database genomic research.

## Results

### JointMR Method overview

Figure 1 provides a visual overview of the JointMR method and our case analysis framework. Consider a scenario where summary-level data are available from *N* independent GWAS databases for an exposure *X* and *M* indpendent GWAS databases for an outcome *Y*. After selecting *J* independent instrumental variables (IVs), we extracted their association estimates (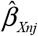 and 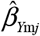) and standard errors (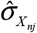 and 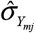) for every *j*-th IV, *n*-th exposure study, and *m*-th outcome study. This *N* × *M* design presents a significant challenge for evidence synthesis. Standard meta-analysis, which often pools MR estimates after they are generated, typically assumes these estimates are independent. However, this assumption is violated: any causal estimates derived using the same outcome database (even with different exposure sources) are inherently correlated because they share the same sampling variation. Failing to account for this correlation structure leads to inefficient estimates, biased standard errors, and potentially incorrect causal inferences. The JointMR framework is proposed to explicitly address this problem. It is a unified joint modeling framework designed to synthesize all *N* × *M* causal estimates simultaneously, while correctly modeling their complex, block-structured covariance.

**Figure 1.**
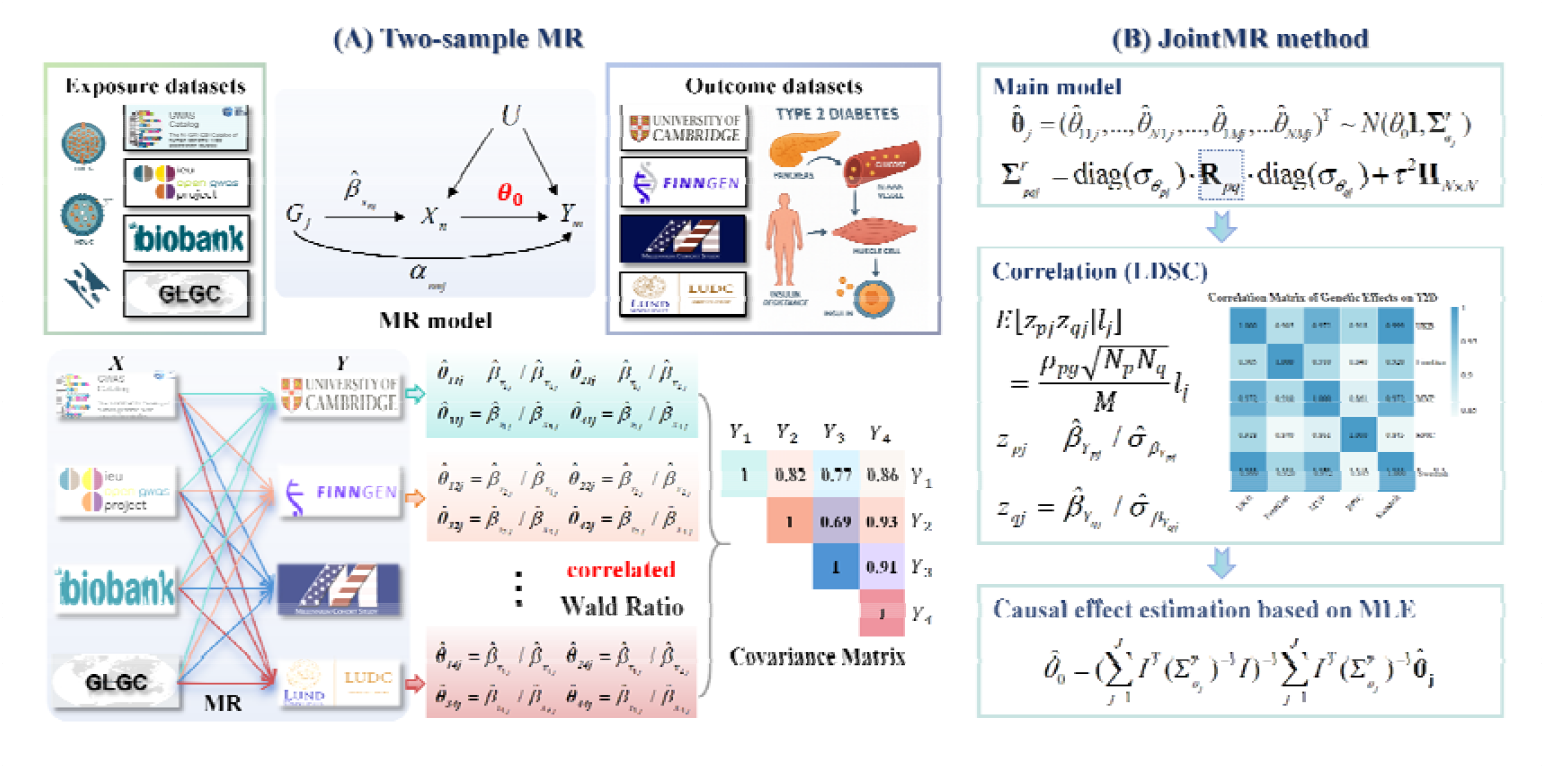
JointMR flowchart. (A) Overview of the two-sample MR framework using SNP–exposure and SNP–outcome summary data. (B) Traditional multi-center MR– meta approach, where study-specific Wald ratios are combined without accounting for cross-study correlations. (C) JointMR integrates correlated MR estimates through a joint covariance structure, providing more efficient and robust causal effect estimation.

We first describe the foundational JointMR Fixed-Effect (JointMR-FE) model, which operates under the three core MR assumptions and additionally assumes a constant causal effect *θ*_0_ across all IVs. The model is built upon the Wald ratio estimates. For each *G*_*j*_, we first calculate the causal effect estimate for every possible *N* × *M* study pairing: 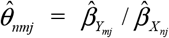, with its variance 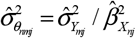 derived using the delta method. This process generates a vector 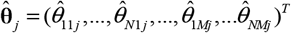 with *N* × *M* causal estimates for each IV. Then for every *G*_*j*_, the JointMR-FE model is defined as:

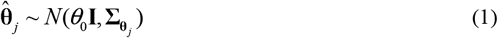

where *θ*_0_ is the true causal effect, and the covariance matrix 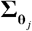 is composed of *M* × *M* blocks and is explicitly modeled as a block matrix to account for the complex dependence structure. The (*p,q*)-th block Σ_*pqj*_ (*p,q*=1,…,*M*) is defined as 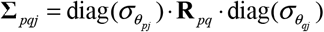, where 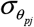 and 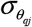 are vectors of standard errors for the causal estimates from the *p*-th and *q*-th outcome studies across *N* exposure studies, and **R**_*pq*_ is an *N*×*N* correlation matrix. Assuming the exposure databases are independent and adhering to the NOME assumption, **R**_*pq*_ captures the correlation structure induced solely by outcome studies *p* and *q*.Then the causal effect 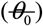 estimation can be obtained by employing the Maximum Likelihood Estimation algorithm.

The JointMR-FE model assumes the exclusion restriction holds, but this is often violated by horizontal pleiotropy, which biases the simple Wald ratios. To address this problem, we extend the JointMR framework by modeling a new JointMR-Wald ratio, which is defined by statistically removing the pleiotropic effect (*α*_*nmj*_) from the outcome association 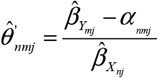. We propose a two-step process to estimate JointMR-Wald ratios for each SNP leveraging MR-Egger regression. Once the corrected ratios 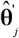 and their corresponding variances are obtained, we substitute them into the model (1) to derive a pleiotropy-robust causal estimate 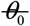.

Additionally, we extend the JointMR-FE model to a random-effects (JointMR-RE) model. This is necessary because in many epidemiological applications, the assumption of a single constant causal effect may be too strong. Different IVs may capture slightly different biological pathways or mechanisms, leading to true heterogeneity in their causal effects, which is distinct from horizontal pleiotropy. The JointMR-RE model accounts for this by assuming the true causal effect *μ*_*j*_ for each IV follows a random distribution around an average causal effect *μ* ~ *N*(*θ*,*τ*^2^), where *τ*^2^ represents the variance of this true heterogeneity. Then we can derive that the marginal distribution for 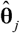 is 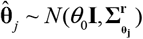, where the (*p,q*)-th block **Σ**^*r*^_*pgj*_ (*p,q*=1,…,*M*) is defined as 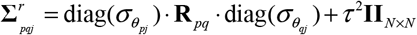. The overall average causal effect 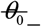 is also jointly estimated using Maximum Likelihood.

### Simulation

We conducted a series of simulation studies to evaluate the performance of the JointMR, comprising eight traditional strategies (formed by applying four MR methods — dIVW [25], fixed/random-effect IVW, and WME — under both “downstream” (MR-meta) and “upstream” (GWAS-meta) frameworks). We assessed variations in the magnitudes of the following parameters in the scenarios of no pleiotropy and horizontal pleiotropy: causal effect, correlation and heterogeneity between databases, the number of databases and the number of SNPs. We utilized boxplots to demonstrate the results of estimation bias and standard error, Q-Q plots to show the results of Type I error, and bar charts to depict the results of statistical power. We also report the empirical coverage of 95% confidence intervals for causal effect estimation, and it is calculated by bootstrap method.

Regarding estimation precision, while all methods produced unbiased estimates centered around zero (Figure 2(A)), JointMR demonstrated superior efficiency. Its variance progressively decreased as the database correlation increased from 0.5 to 0.95, a range chosen to reflect realistic levels of genetic correlation commonly observed among large-scale GWAS consortia and biobank-based datasets. This improvement—unique to our method—confirms that JointMR effectively leverages the correlation structure to enhance precision, a feature absent in the conventional strategies.

**Figure 2.**
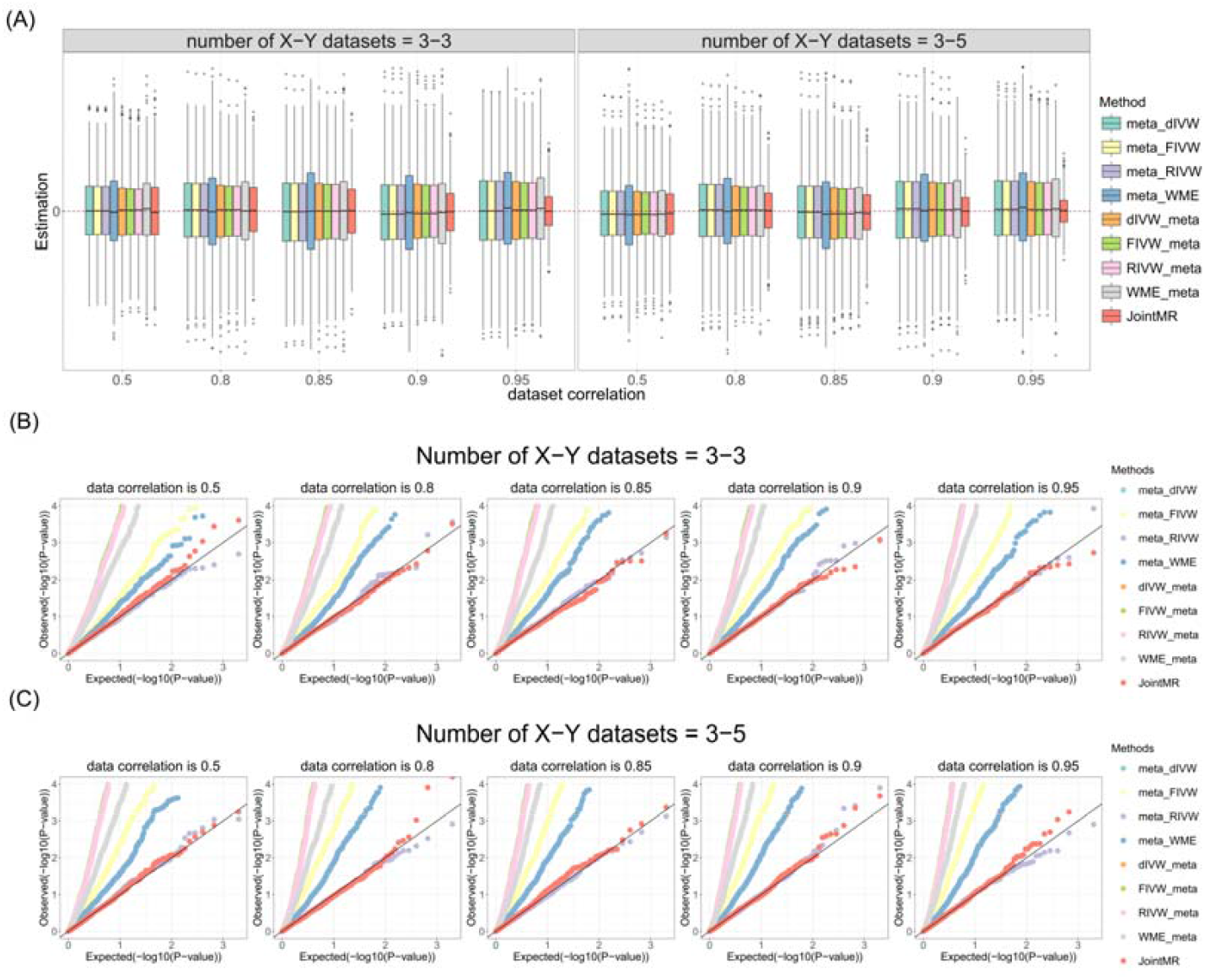
Simulation results for causal effect estimation under the null scenario (no pleiotropy). (A) Boxplots show the performances of causal effect estimation when the true causal effect is zero. and (C) Q-Q plots show the performances of Type I error rates for testing the null causal effect when the number of G-X and G-Y databases is 3 and 3 (B) and 3 and 5 (C), respectively. The eight traditional strategies (legend items) combine four MR methods (dIVW, FIVW, RIVW, WME) with two frameworks: the “meta_” prefix (upstream/GWAS-meta) and the “_meta” prefix (downstream/MR-meta). FIVW, fixed-effect IVW; RIVW, random-effect IVW.

A critical challenge in multi-population MR analysis is the potential inflation of false positives when data correlation is ignored. Under the null hypothesis (true causal effect = 0), as shown in Figure 2, our primary focus was on the validity of Type I error control. The Q-Q plots (Figure 2(B)-(C)) reveal a striking contrast: while JointMR remained robust across all scenarios with P-values adhering closely to the expected uniform distribution, several conventional strategies exhibited noticeable Type I error inflation. Crucially, this inflation in traditional methods was exacerbated as the correlation between databases increased, highlighting their inability to account for shared information. In contrast, JointMR effectively controlled the false positive rate regardless of the correlation structure.

Furthermore, Under the non-null scenario (true causal effect = 0.05, Figure 3), JointMR continued to provide accurate, unbiased estimates (Figure 3(A)). The precision advantage observed under the null hypothesis translated directly into superior statistical power. Figure 3(B)-(C) demonstrates that JointMR was substantially more powerful than all comparator methods across all conditions. Notably, while the power of conventional methods showed only modest gains with increasing database correlation, the power of JointMR increased rapidly. This indicates that our method is uniquely positioned to detect true causal effects in correlated data (Figure S1). As shown in Table 1, JointMR maintained robust 95% confidence interval coverage stable around the nominal 0.95 level across all scenarios, whereas most conventional methods suffered from severe under-coverage, particularly at high database correlations.

**Table 1.** Empirical coverage of 95% confidence intervals for causal effect estimation.

| Database | $\theta_0$ | $\rho_{pq}$ | GWAS-meta | | | | MR-meta | | | | JointMR |
| --- | --- | --- | --- | --- | --- | --- | --- | --- | --- | --- | --- |
|  |  |  | dIVW | fIVW | rIVW | WME | dIVW | fIVW | rIVW | WME |  |
| 3-3 | 0 | 0.3 | 0.903 | 0.903 | 0.958 | 0.901 | 0.658 | 0.658 | 0.958 | 0.722 | 0.961 |
| 3-5 | 0 | 0.3 | 0.852 | 0.851 | 0.951 | 0.887 | 0.595 | 0.595 | 0.951 | 0.676 | 0.943 |
| 3-3 | 0 | 0.9 | 0.773 | 0.773 | 0.935 | 0.845 | 0.500 | 0.500 | 0.935 | 0.610 | 0.936 |
| 3-5 | 0 | 0.9 | 0.661 | 0.661 | 0.959 | 0.771 | 0.416 | 0.416 | 0.959 | 0.510 | 0.936 |
| 3-3 | 0.05 | 0.3 | 0.888 | 0.890 | 0.941 | 0.916 | 0.650 | 0.649 | 0.941 | 0.733 | 0.939 |
| 3-5 | 0.05 | 0.3 | 0.827 | 0.825 | 0.955 | 0.874 | 0.572 | 0.574 | 0.955 | 0.673 | 0.956 |
| 3-3 | 0.05 | 0.9 | 0.758 | 0.757 | 0.945 | 0.839 | 0.519 | 0.519 | 0.945 | 0.607 | 0.946 |
| 3-5 | 0.05 | 0.9 | 0.656 | 0.652 | 0.957 | 0.790 | 0.443 | 0.445 | 0.957 | 0.513 | 0.940 |

**Figure 3.**
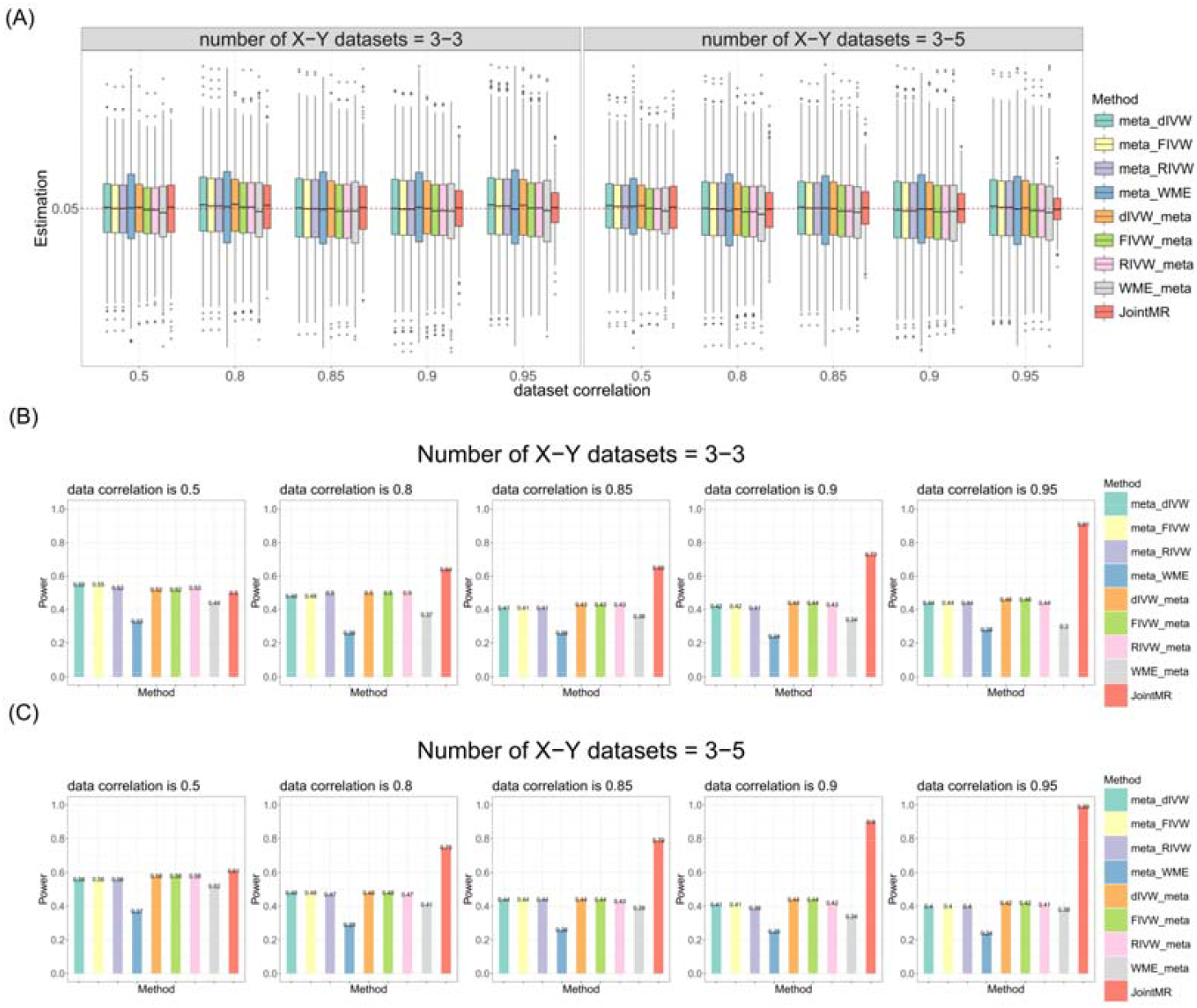
Simulation results for causal effect estimation and statistical power under a non-null scenario (no pleiotropy). (A) Boxplots show the performances of causal effect estimation when the true causal effect is 0.05. (B) and (C) Bar charts show the statistical power for detecting the non-zero causal effect when the number of G-X and G-Y databases is 3 and 3 (B) and 3 and 5 (C), respectively. Method abbreviations are as described in Figure 2.

We then extended our simulations to scenarios where horizontal pleiotropy was present. Under the null hypothesis (Figure 4), the limitations of existing approaches became evident. As expected, conventional methods that do not account for pleiotropy, particularly those based on standard IVW, produced heavily biased causal effect estimates. Notably, even methods typically considered more robust, such as WME, failed to maintain accuracy in these complex multi-population scenarios. In sharp contrast, our JointMR method was the only strategy to provide largely unbiased estimates, successfully mitigating the impact of pleiotropy where all other methods faltered (Figure 4(A)). This disparity was further reflected in the Type I error assessment. The Q-Q plots reveal that the presence of pleiotropy led to substantially inflated Type I error rates across all comparator methods. JointMR, however, demonstrated superior control over Type I error, with its P-value distribution remaining well-calibrated and adhering to the null expectation, unlike the inflated statistics observed in the traditional strategies (Figure 4(B)-(C)).

**Figure 4.**
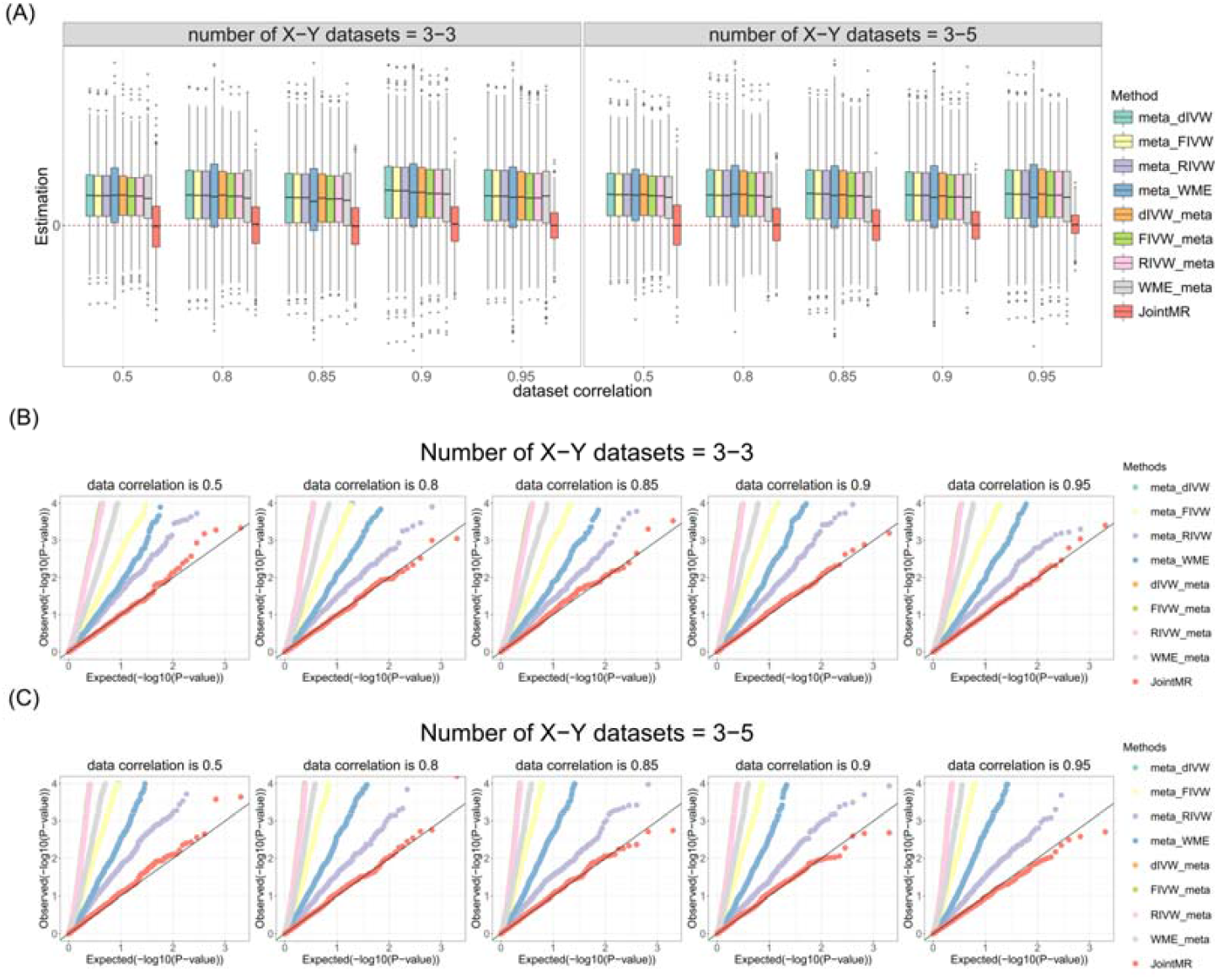
Simulation results for causal effect estimation under the null scenario (horizontal pleiotropy). (A) Boxplots show the performances of causal effect estimation when the true causal effect is zero. (B) and (C) Q-Q plots show the performances of Type I error rates for testing the null causal effect when the number of G-X and G-Y databases is 3 and 3 (B) and 3 and 5 (C), respectively.

Figure 5 demonstrates the performance in the non-null, pleiotropic scenario (true effect = 0.05). Consistent with the null findings, estimates from conventional methods were compromised by bias. JointMR stood out by maintaining accurate, unbiased estimates, successfully resisting the pleiotropic bias that affected all other strategies.

**Figure 5.**
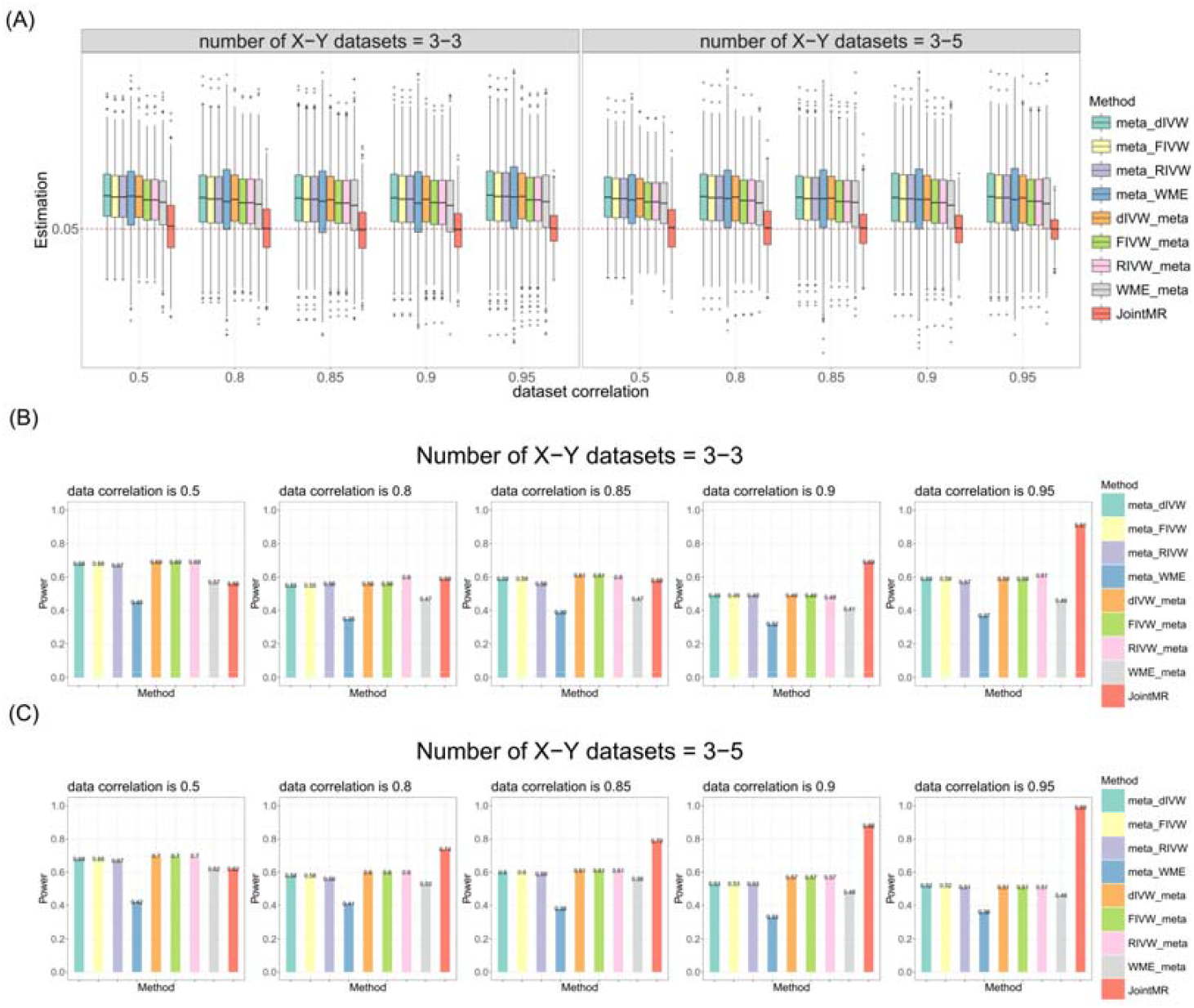
Simulation results for causal effect estimation and statistical power under a non-null scenario (horizontal pleiotropy). (A) Boxplots show the performances of causal effect estimation when the true causal effect is 0.05. (B) and (C) Bar charts show the statistical power for detecting the non-zero causal effect when the number of G-X and G-Y databases is 3 and 3 (B) and 3 and 5 (C), respectively. Method abbreviations are as described in Figure 2.

Furthermore, this robustness did not come at the cost of sensitivity; JointMR exhibited a decisive statistical power advantage over the other methods. This unique combination of unbiased estimation and high statistical power underscores JointMR’s reliability for detecting true causal effects in the presence of horizontal pleiotropy, a scenario where conventional methods proved inadequate (Figure S2).

We re-evaluated the robustness of our method against weak instruments using a revised parameter setting, simulating a stringent scenario where 30% and 50% of instruments were weak. As illustrated in the supplementary results (e.g., Figure S3-S6), JointMR exhibited remarkable stability. The estimation remained unbiased with high precision, and Type I error rates were well-controlled, confirming the method’s reliability even when instrument strength is compromised.

Finally, we assessed performance in the presence of heterogeneity in causal effects across databases. While the introduction of heterogeneity generally increased estimation variance and slightly reduced power for all methods, JointMR remained the most robust approach. Consistent with the homogeneous scenarios, it provided unbiased estimates and controlled Type I errors. Although the gain in precision with increasing correlation was less pronounced than in the fixed-effect model (homogeneous scenario), JointMR still consistently achieved better precision and higher statistical power than all conventional approaches (Figure S7-S12).

### Application

In this section, we applied JointMR to investigate the causal relationships between four major blood lipid traits—high-density lipoprotein cholesterol (HDL-C), low-density lipoprotein cholesterol (LDL-C), total cholesterol (TC) and triglycerides (TG) and the risk of type 2 diabetes (T2D) of European ancestry. For the lipid exposures, data from two distinct databases were analyzed for each trait, sourced from a pool of resources including the UK Biobank (UKB) [26,27], the Global Lipids Genetics Consortium (GLGC) [28], and the Million Veteran Program (MVP) [29]. For the T2D outcome, summary statistics were incorporated from five databases: the UKB [30], FinnGen, MVP, the EPIC-InterAct project (EPIC) [31], and the Swedish ANDIS [32] (Table S1). We compare JointMR’s results directly against two conventional summary-level strategies: GWAS-meta MR (corresponding to the GWAS-meta strategy) and traditional MR-meta (corresponding to the MR-meta strategy). The results presented for these conventional strategies are based on the widely-used random-effects IVW method.

We first estimated the correlation structure of the genetic effects on T2D risk across the five outcome GWAS databases. The analysis revealed a moderate and heterogeneous correlation structure, with correlation coefficients ranging from 0.840 to 0.999. This indicates strong correlations between the databases, as they share significant underlying genetic signals (Figure 6).

**Figure 6.**
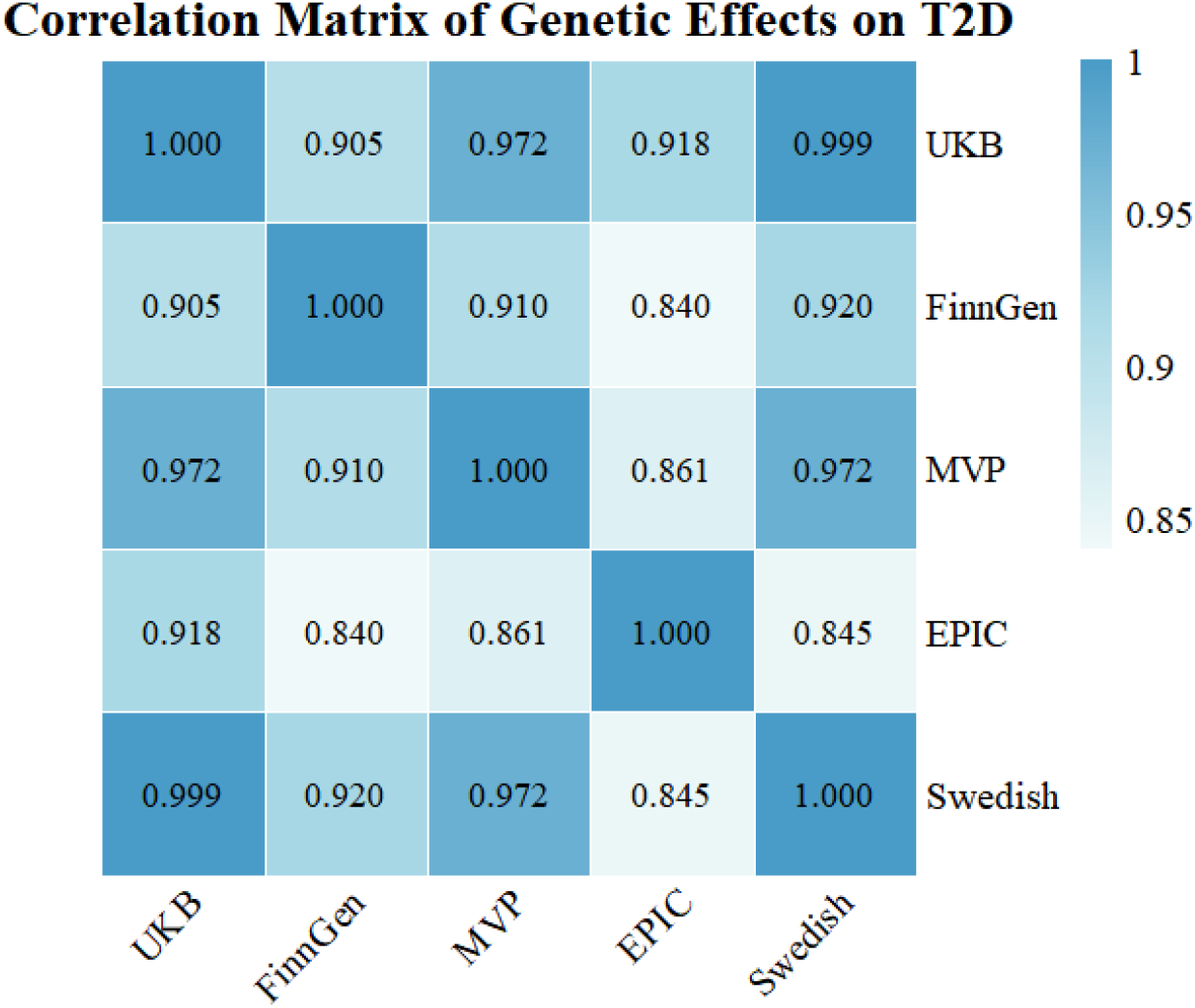
Correlation heatmap of genetic effects on T2D. The heatmap visualizes the estimated correlation matrix of genetic effects on T2D risk across the five outcome GWAS databases.

The analysis of TC revealed a stark contradiction between the two conventional strategies (Figure 7(A)). The GWAS-meta strategy identified a minor but significant risk effect (*OR* = 1.006, *P* = 0.013), while the MR-meta strategy suggested a significant protective effect (*OR* = 0.911, *P* < 1.425×10^−4^). This fundamental disagreement creates significant ambiguity regarding the true causal direction. JointMR resolves this conflict by integrating all data sources simultaneously. It not only refutes the protective finding but also identifies a much stronger, highly significant risk effect (*OR* = 1.236, *P* = 8.212×10^−20^) that was obscured by the simpler methods. This robust finding aligns with recent MR studies identifying TC as a risk factor for T2D [33–34] and demonstrates JointMR’s ability to overcome the instability that confounded the traditional approaches.

**Figure 7.**
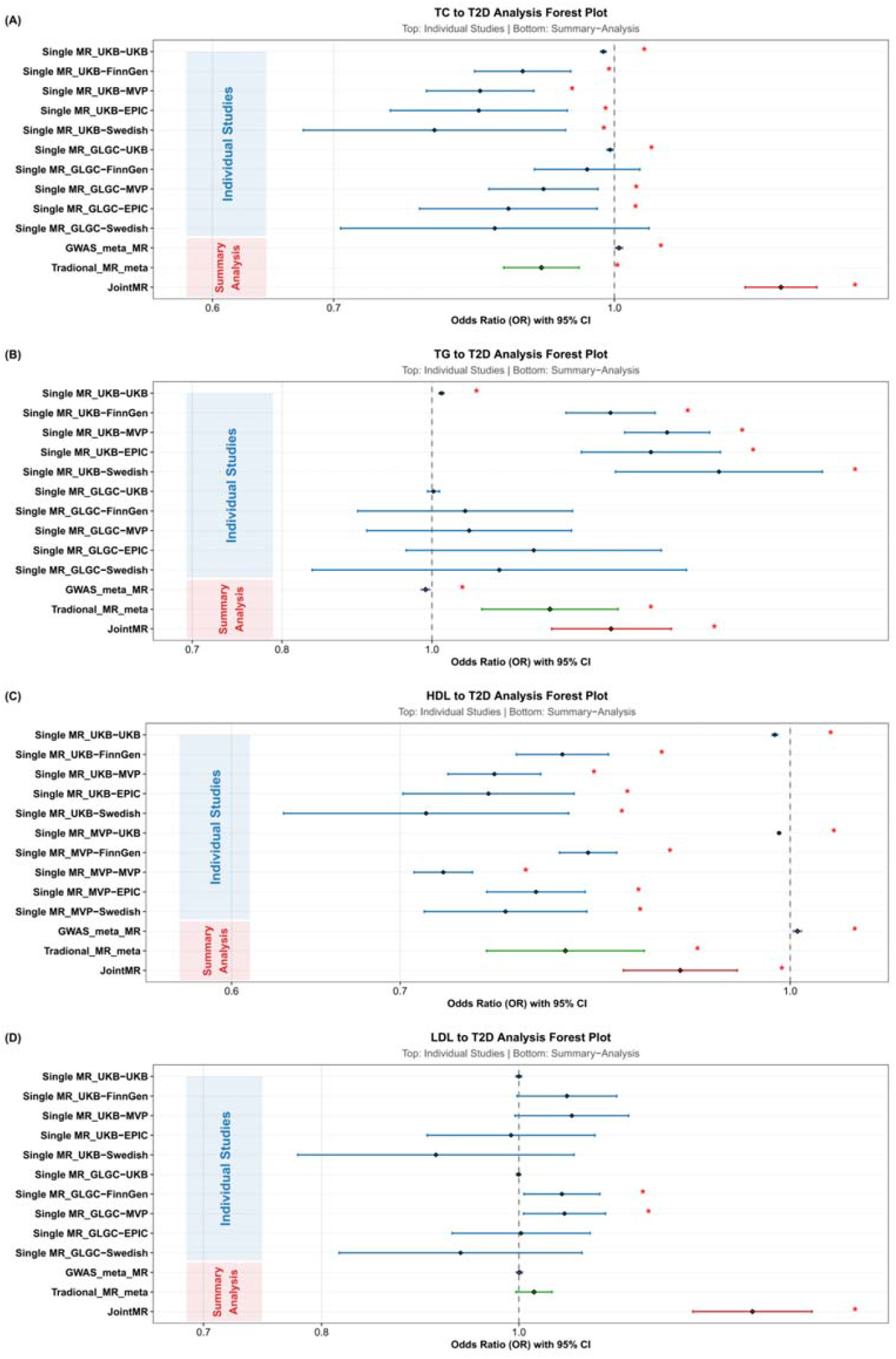
Forest plots displaying ORs with 95% confidence intervals. The plot showed the causal effects of (A) TC, (B) TG, (C) HDL-C and (D) LDL-C on T2D risk. Each plot compares results from individual two-sample MR analyses (upper section, blue background) against summary estimates from three strategies (lower section, red background). Individual studies are labeled following the convention “Single MR_[Exposure database]-[Outcome database]” representing specific database pairings. The summary methods include GWAS-meta followed by MR (green), traditional MR meta-analysis (purple), and the novel JointMR framework (red) that explicitly accounts for between-study correlations. The vertical dashed line indicates the null effect (*OR* = 1). Red asterisks denote statistically significant results (*P* < 0.05). The JointMR method demonstrates enhanced stability and precision compared to conventional approaches, particularly when integrating evidence from multiple data sources with correlated structures.

For TG, the discrepancy between conventional approaches was particularly notable (Figure 7(B)). The GWAS-meta approach indicated a significant protective effect (*OR* = 0.991, *P* = 1.845×10^−3^), whereas the MR-meta approach suggested a significant risk effect (*OR* = 1.192, *P* = 6.893×10^−4^), producing completely opposite conclusions. This fundamental disagreement highlights their instability when faced with significant data heterogeneity. JointMR effectively resolved this critical ambiguity, integrating the conflicting evidence to identify a clear and significant risk effect (*OR* = 1.306, *P* = 6.379×10^−7^). This robust finding, which aligns with recent metabolomic studies [35] and reflects the complex nature of the TG-T2D link [36], demonstrates JointMR’s superior stability in neutralizing conflicting data into a single, coherent estimate.

For HDL-C, conventional methods yielded inconsistent conclusions regarding a causal effect (Figure 7(C)). The GWAS-meta strategy produced a near-null result with a minimal risk estimate (*OR* = 1.007, *P* = 4.620×10^−4^), whereas the MR-meta approach suggested a significant protective effect (*OR* = 0.814, *P* = 2.276×10^−8^). In contrast, JointMR identified a clear and robust protective effect of HDL-C (*OR* = 0.904, *P* = 1.536×10^−4^). This coherent and statistically robust effect, supported by prior genetic and epidemiological evidence on HDL-C [37–38], highlights JointMR’s ability to reconcile heterogeneous and seemingly contradictory MR signals.

JointMR’s superior efficacy was further confirmed in the analysis of LDL-C (Figure 7(D)). Both conventional strategies failed to establish a significant causal relationship, with their confidence intervals crossing 1. Only JointMR identified a clear and statistically significant risk-increasing effect (*OR* = 1.302, *P* = 1.509×10^−14^). This finding robustly reinforces the established biological consensus that genetically elevated LDL-C is a causal risk factor for T2D, a link overwhelmingly supported by a vast body of MR literature, including recent large-scale studies [39–41]. The failure of conventional methods to confirm this well-known, clinically relevant association demonstrates their limited statistical power and further validates JointMR’s reliability in identifying true causal effects.

## Discussion

In this study, we developed JointMR, a likelihood-based meta-method designed for integrating multiple GWAS summary datasets to obtain more powerful and reliable MR effect estimates. Through comprehensive simulations and real-data applications, we demonstrate that JointMR markedly outperforms the two most widely used conventional strategies, GWAS-meta and MR-meta. Across all scenarios, JointMR delivers higher statistical power, well-controlled Type I error rates, and, importantly, substantially reduced variance inflation, a known problem in multi-cohort MR that often leads to unstable or misleading causal conclusions.

Our real-world lipids-T2D application further illustrates these advantages. In the TC, TG and HDL-C analysis, the two traditional approaches produced directly contradictory results, reflecting their sensitivity to heterogeneous and conflicting databases. JointMR effectively resolved this inconsistency, yielding a stable and biologically plausible near-null estimate. Likewise, only JointMR had sufficient statistical power to detect the risk effect of LDL-C, while maintaining variance stability and proper error control. These results underscore JointMR’s ability to extract maximal information from multiple correlated datasets without amplifying noise, an essential capability in modern MR studies where database overlap and sample correlation are unavoidable.

Methodologically, JointMR improves upon existing approaches in two important dimensions. First, unlike the GWAS-meta strategy, which collapses exposures and outcomes into aggregated averages, JointMR directly models the full joint likelihood and covariance structure across all exposure–outcome pairs, avoiding aggregation-induced biases such as Simpson’s paradox [42]. Second, unlike MR-meta, which analyzes each pair in isolation and pools afterward, JointMR borrows strength across all databases in a unified model, yielding more precise and robust causal estimates. This joint modeling framework also implicitly reduces the impact of publication bias and selective database inclusion, as each dataset contributes proportionally to its information content rather than disproportionately influencing pooled results. Our simulation results provide additional practical guidance: the precision benefits of JointMR increase substantially when integrating highly correlated or overlapping databases, exactly the setting in which conventional meta-strategies tend to suffer the most severe variance inflation and Type I error inflation. This highlights the suitability of JointMR for modern biobank-scale consortia, where overlapping samples and correlated designs are the norm rather than exceptions.

Finally, the random-effects extension (JointMR-RE) plays a critical role in real-world applications. By allowing the causal effect to vary across databases, JointMR-RE quantifies between-database causal heterogeneity, a form of heterogeneity fundamentally different from pleiotropy. Even within European ancestry cohorts, subtle differences in recruitment schemes, environmental backgrounds, and fine-scale population structures can induce genuine effect variation. JointMR-RE captures these variations without sacrificing stability or inflating false positives. This feature also positions JointMR as a strong candidate for future trans-ethnic MR analyses, where modeling causal heterogeneity is essential.

However, our study has several limitations. First, while JointMR accounts for directional horizontal pleiotropy, it does not explicitly model or correct for correlated pleiotropy or vertical pleiotropy [43]. Second, our method relies on the NOME assumption for the SNP-exposure effects, and it does not account for potential sample overlap between the exposure databases themselves, or between the exposure and outcome databases, which can introduce bias [44]. Third, our application was restricted to European ancestry cohorts, the method’s performance in trans-ethnic analyses is complicated by differing LD structures, which remains to be evaluated. Finally, the process of harmonizing SNPs across all studies requires taking an intersection, which can lead to a small number of shared instruments, increasing the risk of weak instrument bias [45].

In conclusion, our proposed JointMR method overcomes critical weaknesses of power and instability that are prevalent in conventional strategies for meta-analyzing MR summary data. By resolving contradictory findings and providing the statistical power to detect subtle causal effects that other methods miss, JointMR represents a more robust and reliable tool for dissecting complex causal relationships from the wealth of available GWAS summary statistics.

## Methods

Consider a scenario where summary-level data are available from *N* independent GWAS databases for an exposure *X* and *M* indpendent GWAS databases for an outcome *Y*. Since we are mainly interested in improving the statistical power of causal effect estimation by integrating these multiple data sources, we chose *J* independent IVs (SNPs) associated with at least one exposure database in all databases. For *j*-th IV (*G*_*j*_), we extract its estimated effect on the exposure from the *n*-th study 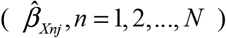 and its effect on the outcome from the *m*-th study 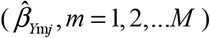, along with their respective standard errors (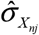 and 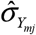).

For each of the *N* × *M* possible pairings of exposure and outcome studies, a causal effect for the *j*-th IV using the Wald ratio is calculated. This estimator relies on the NOME assumption, meaning the SNP-exposure effect is estimated without error. The Wald ratio is 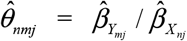,with its variance using the delta method, 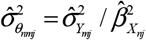. We subsequently define the vector of estimated causal effects for the *j*-th IV, collating all study pairings, as 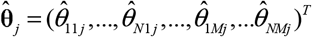.

### JointMR based on fixed-effect model

Our primary goal is to estimate the single, true causal effect (*θ*_0_) of *X* on *Y*, assuming this effect is constant across all studies and IVs. This model relies on each IV *G*_*j*_ satisfying the three core MR assumptions: (1) Relevance: The IV is associated with the exposure *X*; (2) Independence: The IV is independent of any confounders of the *X*-*Y* relationship; (3) Exclusion Restriction: The IV affects the outcome *Y* only through the exposure *X*.

Based on these assumptions, the vector of true causal effects for the *j*-th IV, 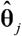, is defined as a vector where each element is the single, constant causal effect *θ*_0_. The corresponding vector of estimated causal effects, 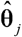, is then modeled as following a multivariate normal distribution centered on this true vector:

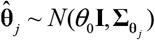

where the covariance matrix 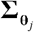 is explicitly modeled as a block matrix to account for the complex dependence structure. This *NM* × *NM* matrix is composed of *M* × *M* blocks, where the (*p,q*)-th block **Σ**_*pqj*_ (*p,q*=1,…,*M*) is defined as:

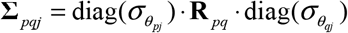

where 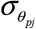 and 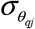 are vectors of standard errors for the causal estimates from the *p*-th and *q*-th outcome studies across *N* exposure studies. **R**_*pq*_ is an *N*×*N* correlation matrix capturing the correlation structure between outcome studies, when estimates share the same outcome database, their correlation simplifies to *ρ* = 1. For estimates from different outcome databases *p* and *q*, the correlation is given by *ρ* = *ρ*_*pq*_, where *ρ*_*pq*_ is the correlation between the z-scores of the genetic associations in the two outcome studies.

By employing the Maximum Likelihood Estimation method, we can obtain the log-likelihood function of model (1)

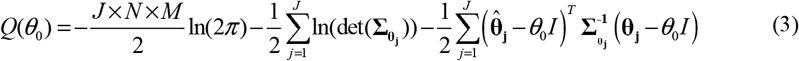

We aimed to maximize the log-likelihood function using the Nelder-Mead method [46] to obtain the estimation of 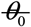. Then, we used the likelihood ratio test to perform hypothesis testing:

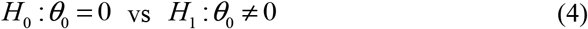

with the testing statistics

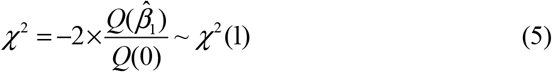

When there is horizontal pleiotropy, the third assumption of MR is violated, the traditional Wald ratio is biased and we model the JointMR-Wald ratio as following

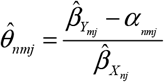

where *α*_*nmj*_ represent the horizontal pleiotropy and it is unknown. Therefore, in the first step, we need to estimate *α*_*nmj*_ using MR-Egger regression

(1) estimate 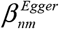 using MR-Egger regression

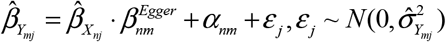
(2) estimate using

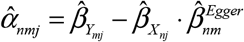

Then we can obtain the estimations of new Wald ratio 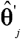 by substituting *α*_*nmj*_ into equation (1), then obtain the estimation of 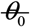.

### JointMR based on random-effect model

To account for potential true heterogeneity across IVs — where different instruments may reflect distinct biological pathways—we extend the JointMR-FE model to a random-effects (JointMR-RE) framework. This model relaxes the fixed-effect assumption of a single constant causal effect *θ*_0_.

Instead, it posits that the true causal effect for the *j*-th IV, *μ*_*j*_, is itself a random variable drawn from a distribution centered around an average causal effect *μ*_*j*_ ~ *N*(*θ*_0_,*τ*^2^). Here, *τ*^2^ represents the variance of this true heterogeneity, which is first estimated using the DerSimonian-Laird method [47]. Incorporating this additional variance component, the marginal distribution for the estimated vector 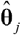 becomes:

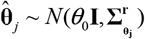

where the (*p,q*)-th block **Σ**^*r*^_*pgj*_ (*p,q*=1,…,*M*) is defined as:

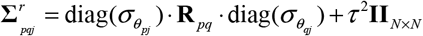

The overall average causal effect 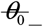 is also is then estimated using Maximum Likelihood based on this marginal distribution.

### Simulation settings

In our simulation study, we systematically evaluated the performance of our proposed method against several conventional approaches. We began by generating GWAS summary statistics that reflect a variety of complex, realistic scenarios for *J* independent SNPs across *N* exposure databases and *M* outcome databases. To realistically generate this summary-level exposure data, we first simulated individual-level genotype and phenotype data for *N*_*1*_ individuals. For each study *n*, the genotype *G*_*nlj*_ for individual *l* and SNP *j* was drawn from a binomial distribution:

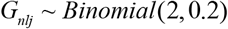

This process allowed us to control for a shared set of true genetic effects 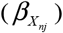 with pre-specified proportions of weak instruments. The continuous exposure phenotype *X*_*nl*_ for individual *l* was then generated based on the true effects:

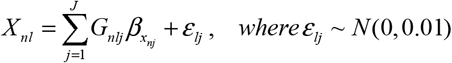

We then mimicked a real GWAS by performing *J* separate linear regressions (*X*_*nl*_ ~ *G*_*nlj*_) for each of the studies. From these regression outputs, we extracted the required summary statistics: the estimated effect size 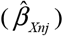, its standard error 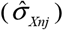 and its P-value. Summary statistics for the *M* outcome databases were generated subsequently based on this foundational exposure data.

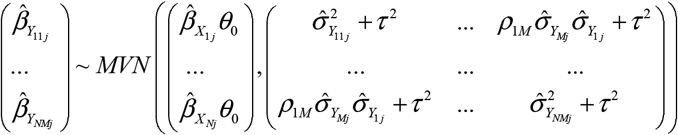

where 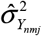 was the variance of 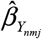. Then we generated the Wald ratios 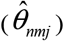 for different databases from above GWAS summary statistics:

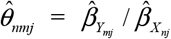

We considered scenarios where the causal effects were either zero 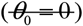 to assess Type I error or nonzero 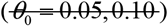 to assess bias and power. We also explored a wide range of scenarios by varying the heterogeneity between studies with 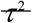 set to 0 and 0.2, and by setting the correlation between GWAS databases *ρ*_*pq*_ vary from 0.5 to 0.95 reflecting correlations observed in real data. To assess the impact of varying study counts, the number of exposure (*N*) databases was fixed at 3, while the number of outcome (*M*) databases was set to 3 and 5. Furthermore, to evaluate the impact of weak instruments, we simulated scenarios by setting the proportion of weak instruments to 0%, 30%, and 50%. Finally, our simulation study was designed to encompass two distinct scenarios: one where pleiotropy was absent (*α*_*nmj*_ = 0), another where directional horizontal pleiotropy was present (*α*_*nmj*_ ~ *U* (0, 0.01)).

To benchmark the performance of our approach, we conducted a comparative analysis with 8 conventional strategies. These strategies were categorized into two distinct approaches, corresponding to the “downstream” and “upstream” practices described earlier, each applying a different IV selection strategy to mitigate weak instrument bias. The “upstream” strategy involved first meta-analyzing the GWAS summary statistics. The resulting SNPs were then filtered to retain only those meeting *P* < 5× 10^−8^ and *F* > 10, after which the four MR methods (dIVW, fixed/random-effect IVW, and WME) were applied. The “downstream” strategy involved initially performing individual MR analyses. For each individual MR, the instrument set was strictly filtered to include only SNPs satisfying *P* < 5× 10^−8^ and *F* > 10 within that specific study, and the resulting causal estimates were then pooled. In contrast, our JointMR approach used a robust selection criteria designed to maximize power while avoiding weak instruments, retaining SNPs that satisfied *F* > 10 in all exposure databases and *P* < 5 × 10^−8^ in at least one exposure database. Finally, to comprehensively evaluate the impact of weak instruments, we varied the proportion of weak instruments ranging from 0 to 0.5 in our simulations.

The evaluation metrics include estimation bias, standard error, type I error for testing null causal effects and statistical power for testing nonnull causal effects. We utilized boxplots to demonstrate the results of bias and standard error, Q-Q plots to showcase the results of Type I error, and bar charts to depict the results of statistical power. We also report the empirical coverage of 95% confidence intervals for causal effect estimation, and it is calculated by bootstrap method.

### Application

To demonstrate the practical utility of our method, we applied it to investigate the causal relationships between four major blood lipid traits—HDL-C, LDL-C, TC, TG and the risk of T2D in individuals of European ancestry. We obtained publicly available GWAS summary statistics from several large-scale consortia. For the lipid traits (HDL, LDL, TC, TG), we utilized summary data from sources including the GLGC, the UKB and the MVP. For the outcome, T2D, summary statistics were sourced from the UKB, the MVP, the FinnGen, the EPIC and the Swedish ANDIS. All databases were based on populations of European descent. The detailed information for each GWAS database is provided in Supplementary Table S1.

This key correlation is estimated from the summary-level data. Specifically, to calculate the genetic correlation (*ρ*_*pq*_) between two outcome studies *p* and *q*, we employed cross-trait LD Score Regression (LDSC). This method leverages genome-wide summary statistics from both studies, rather than only the pre-selected IVs. By regressing the product of Z-scores against a pre-computed LD reference panel, the method accurately estimates *ρ*_*pq*_ while accounting for linkage disequilibrium (LD) and potential sample overlap [48]. The detailed formula is provided in the Appendix (See Supplementary Note).

For each exposure, we firstly obtained the union set of significant SNPs in all databases of exposure. Then we calculated the combined P-value of all databases for each SNP in the union set. The combined P-value for *j*-th SNP was calculated by the Fisher’s method [49–50]: *p*_*j*_ = 2× (1− Φ(*Z*_*j*_)), where *Z*_*j*_ = ∑_*e*_ *Z*_*je*_ = ∑_*e*_ Φ^−1^(1− *p*_*je*_ / 2), Φ was the standard normal cumulative distribution function and *p*_*je*_ was the P-value for *j*-th SNP in *e*-th database. Finally, we conducted the linkage disequilibrium clump process (*r*^*2*^ < 0.001, window size = 1000 kb) based on the combined P-value using the 1000 Genomes Project European reference panel to ensure independence. For two SNPs with high linkage disequilibrium, we kept the SNP with smaller combined P-value. Using the selected instruments for each exposure, we applied our proposed method, alongside the eight conventional strategies described previously, to estimate the causal effect of each lipid marker on T2D.

## Acknowledgments

We thank all investigators and participants of the FinnGen study, UK Biobank and the Department of Veterans Affairs (VA) Million Veteran Program (MVP). We are grateful to the supports of the National Natural Science Foundation of China, the National Key Research and Development Program of China, the Shandong Provincial Natural Science Foundation of China, and Shandong Provincial Key Research and Development project.

## Data Availability Statement

The GWAS summary data in the FinnGen are publicly available at FinnGen (R12 release): https://www.finngen.fi. Other GWAS summary data are publicly available at the GWAS Catalog: https://www.ebi.ac.uk/gwas/. All the analyses in our article were implemented by R software (v.4.3.2). R packages used in our analysis include ldscr, MendelianRandomization, TwoSampleMR, ggplot2, plinkbinr, mr.divw, meta and ieugwasr. The JointMR package can be implemented by GitHub: https://github.com/hhoulei/JointMR. All the codes for simulation are uploaded in GitHub: https://github.com/hhoulei/JointMR_Simul.

## Author Contributions

HLand FX conceived the study. SW and LH contributed to theoretical derivation. SW contributed to the data simulation. LH and SW contributed to the application. LH and SW wrote the manuscript. ZY, XS, YY, HC and LH participated in the discussion and review of the manuscript. All authors reviewed and approved the final manuscript.

## Supplementary File

### Supplementary Materials and Methods

Details of methods and results of simulation.

### Supplementary Figures

Simulation results

### Supplementary Table

GWAS summary databases information and results of application.

