## Supplemental File for "JointMR: A joint likelihood-based approach for causal effect estimation in overlapping Mendelian Randomization studies"

**Supplementary Materials and Methods**

**Theoretical derivation and methodology details**

***Fixed-effect model***

For every
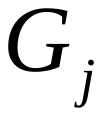
, the vector of causal effect estimates across all *N* × *M* study pairings is modeled as:

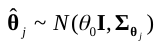

where,
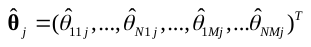
 represents the vector of causal estimates from the *M* outcome database across the *N* exposure studies. The covariance matrix
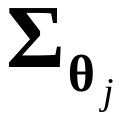
 is explicitly modeled as a block matrix to account for the complex dependence structure. This *NM* × *NM* matrix is composed of *M* × *M* blocks, where the (*p*,*q*)-th block
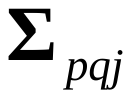
 (*p*,*q*=1,…,*M*) is defined as:

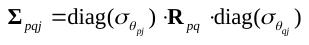

where
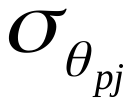
 and
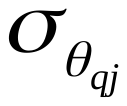
 are vectors of standard errors for the causal estimates from the *p*-th and *q*-th outcome studies across *N* exposure studies.
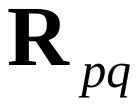
 is an *N*×*N* correlation matrix capturing the correlation structure between outcome studies, when estimates share the same outcome dataset, their correlation is

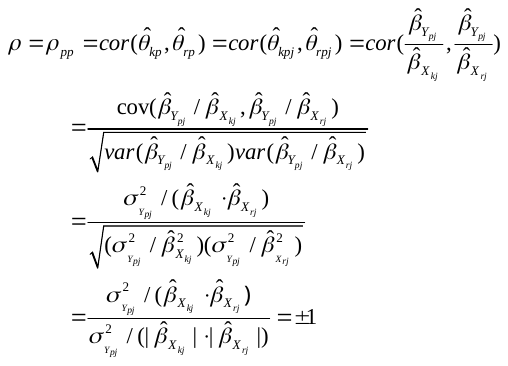

In our framework, we constrain this correlation to 1, based on the biological assumption that the direction of the SNP-exposure effect is consistent across studies. For estimates from different outcome datasets *p* and *q*, the correlation is given by

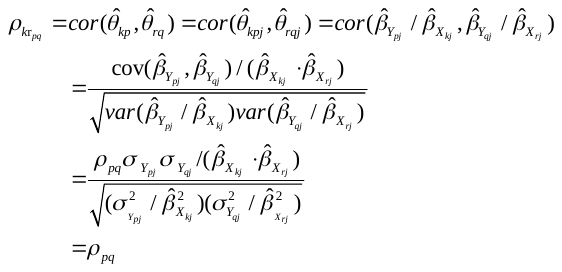

To estimate the genetic correlation
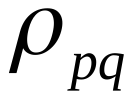
 between two studies *p* and *q*, LDSC models the expected product of their Z-scores (
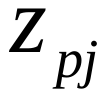
 and
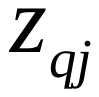
) as a function of the LD score
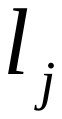
:

$E[z_{pj}z_{qj}|l_{j}]=\frac{\rho_{pg}\sqrt{N_{p}N_{q}}}{J}l_{j}+\frac{\rho_{s}N_{s}}{\sqrt{N_{p}N_{q}}},$
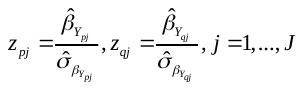

where
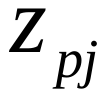
 and
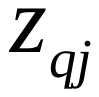
 are the Z-scores for SNP *j* from studies *p* and *q*, $N_{p}$ and $N_{q}$ are their respective sample sizes, $l_{j}$ is the LD score of *jth* SNP, $N_{s}$ is the number of overlapping samples,
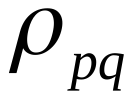
 is the correlation between the two databases and $\rho$ is the confounding covariance caused by sample overlap. The ldscr R package performs this regression by regressing the product
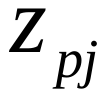
 and
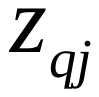
 onto
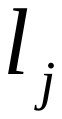
 across all genome-wide SNPs. It uses the slope to estimate the genetic covariance and then standardizes this value using the single-trait heritability estimates to provide the final genetic correlation
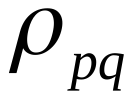
.

The log-likelihood function is

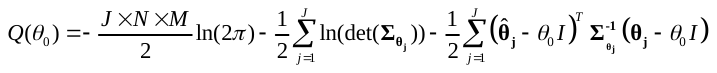

Finally, we obtain the solution of
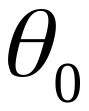

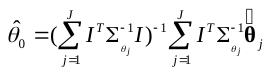

***Random-effect model***

Furthermore, if we assume the causal effect
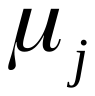
 is not constant but follows a random effects model, i.e.
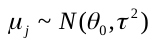
, then our model becomes:

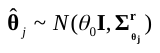

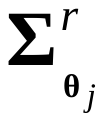
 is also composed of *M* × *M* blocks, where the (*p*,*q*)-th block
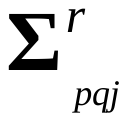
 (*p*,*q*=1,…,*M*) is defined as:

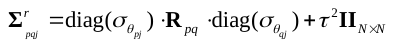

where
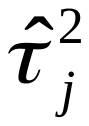
 is calculated using the DerSimonian-Laird method.

Then, we obtained the estimation of
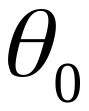
 via maximum likelihood estimation using the Nelder-Mead method.

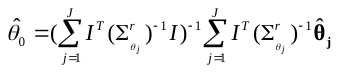

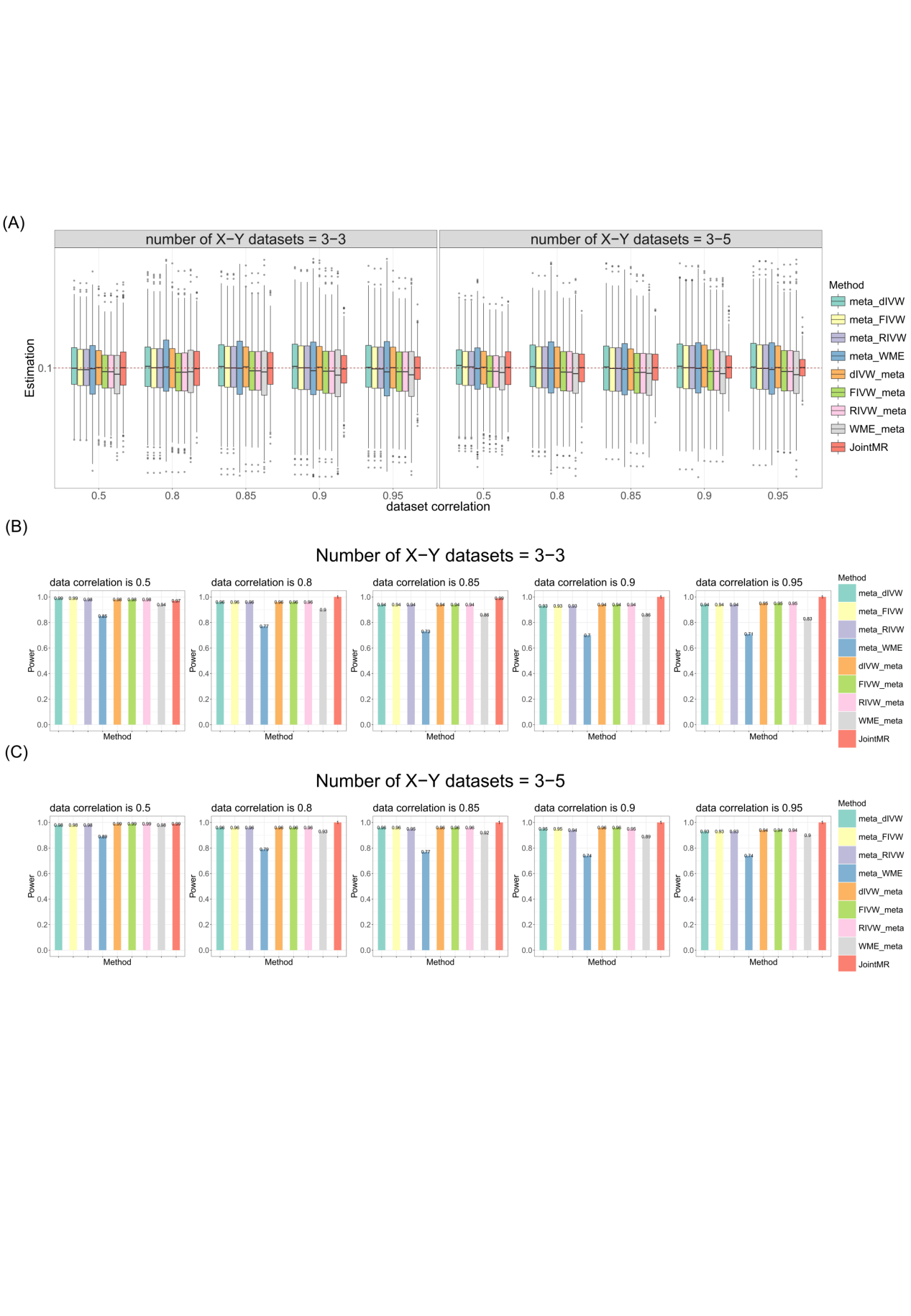

**Figure S1. Simulation results for causal effect estimation under the null scenario (causal effect =0.1, no pleiotropy).**

(A) Boxplots show the performances of causal effect estimation when the true causal effect is 0.1. (B) and (C) Bar charts show the statistical power for detecting the non-zero causal effect when the number of G-X and G-Y databases is 3 and 3 (B) and 3 and 5 (C), respectively.

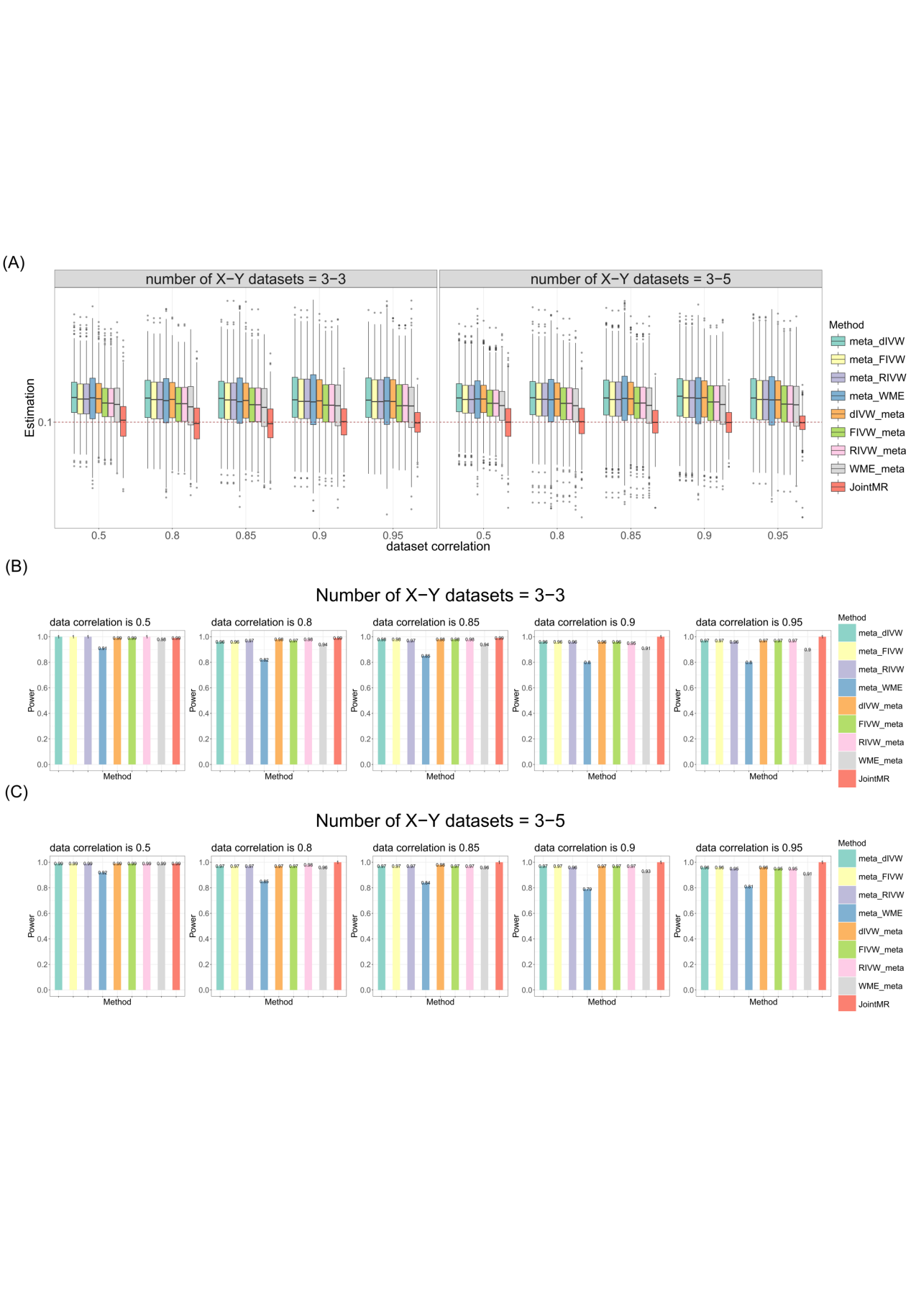

**Figure S2. Simulation results for causal effect estimation under the null scenario (causal effect =0.1, horizontal pleiotropy).**

(A) Boxplots show the performances of causal effect estimation when the true causal effect is 0.1. (B) and (C) Bar charts show the statistical power for detecting the non-zero causal effect when the number of G-X and G-Y databases is 3 and 3 (B) and 3 and 5 (C), respectively. Method abbreviations are as described in Figure 2.

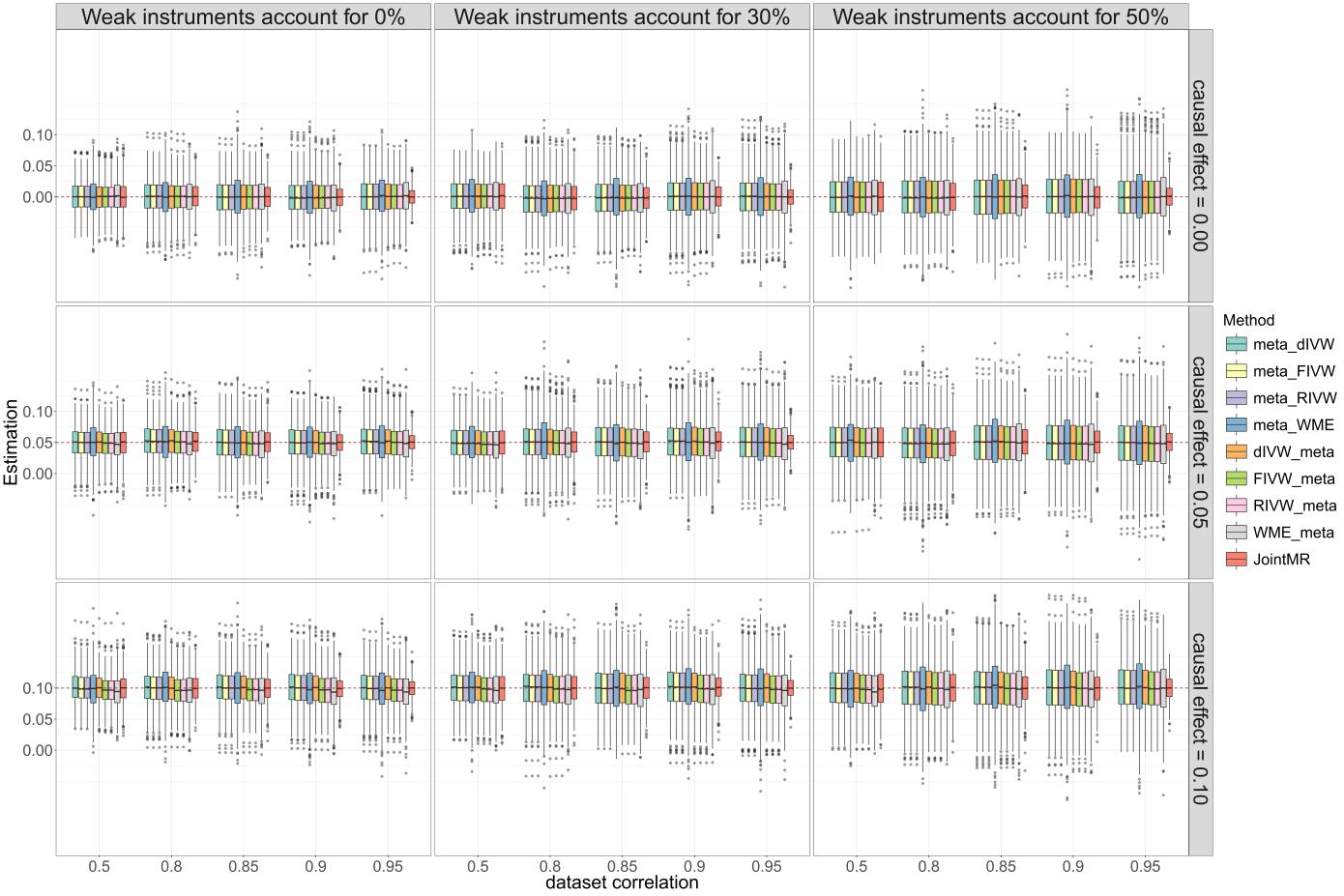

**Figure S3. Simulation results for causal effect estimation by proportion of weak instruments.**

Boxplots show the performances of causal effect estimation across a range of true causal effects (columns), numbers of SNPs (rows), and dataset correlations (x-axis within each panel). The dashed line in each panel indicates the true causal effect.

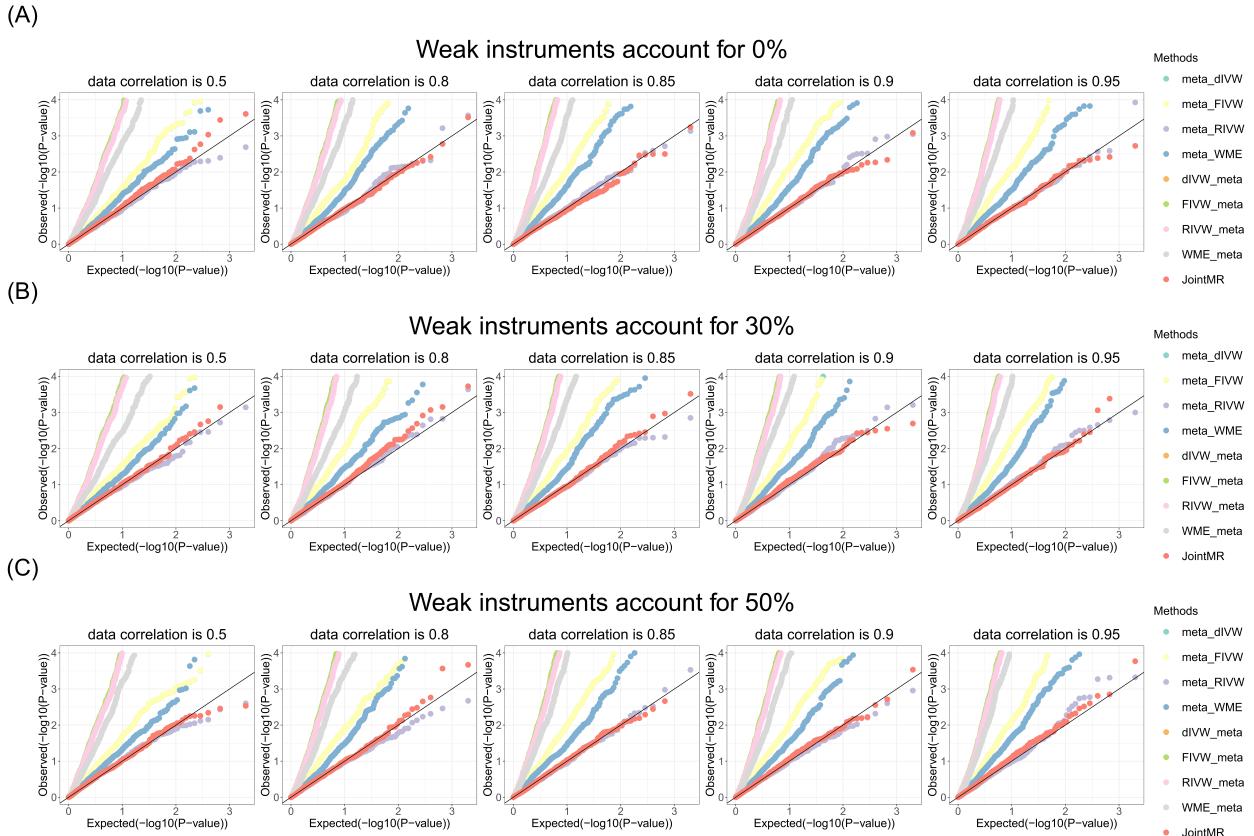

**Figure S4. Simulation results for Type I error rates under varying proportions of weak instruments.**

Q-Q plots show the performances of Type I error rates for testing the null causal effect under scenarios with varying proportions of weak instruments: (A) 0%, (B) 30%, and (C) 50%. The diagonal line represents the expected uniform distribution of P-values under the null hypothesis.

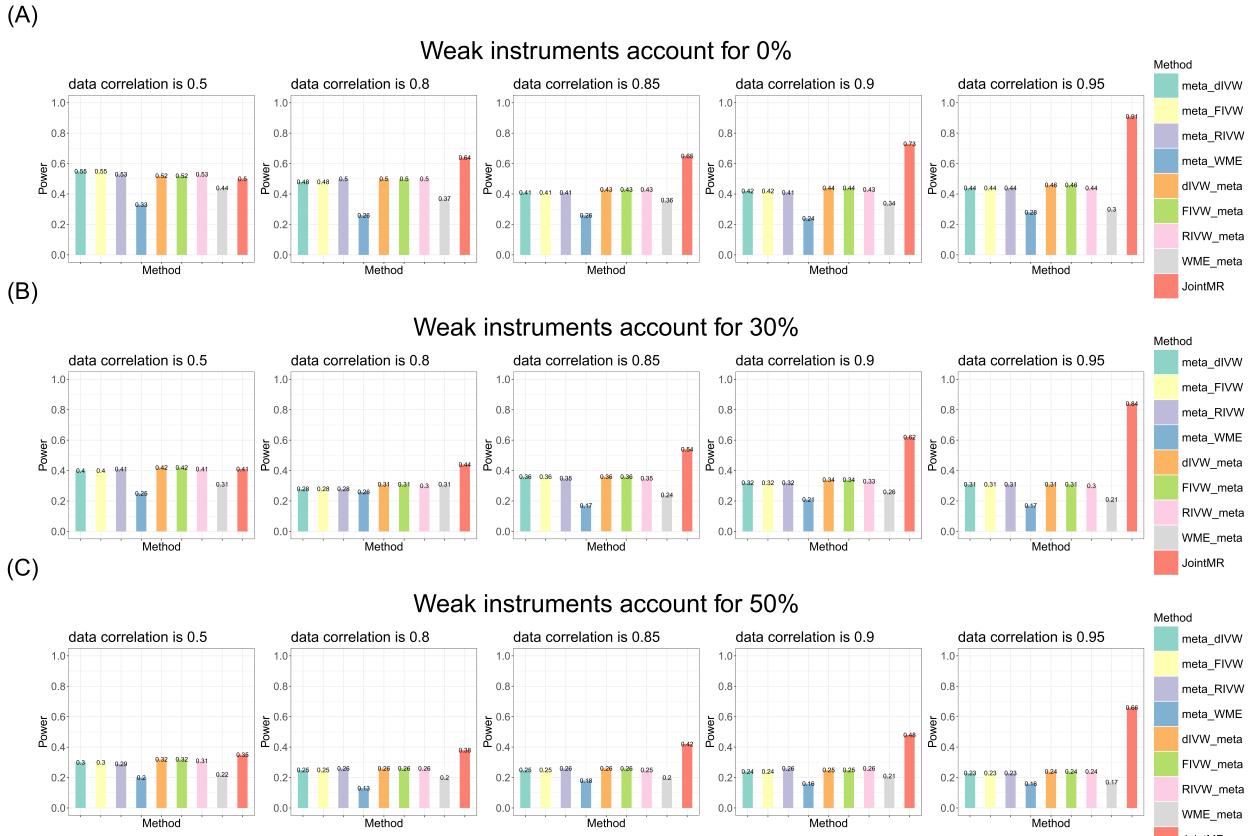

**Figure S5. Simulation results for statistical power by proportion of weak instruments (causal effect = 0.05).**

Bar charts show the statistical power for detecting a non-zero causal effect (true effect = 0.05) under scenarios with varying proportions of weak instruments: (A) 0%, (B) 30%, and (C) 50%.

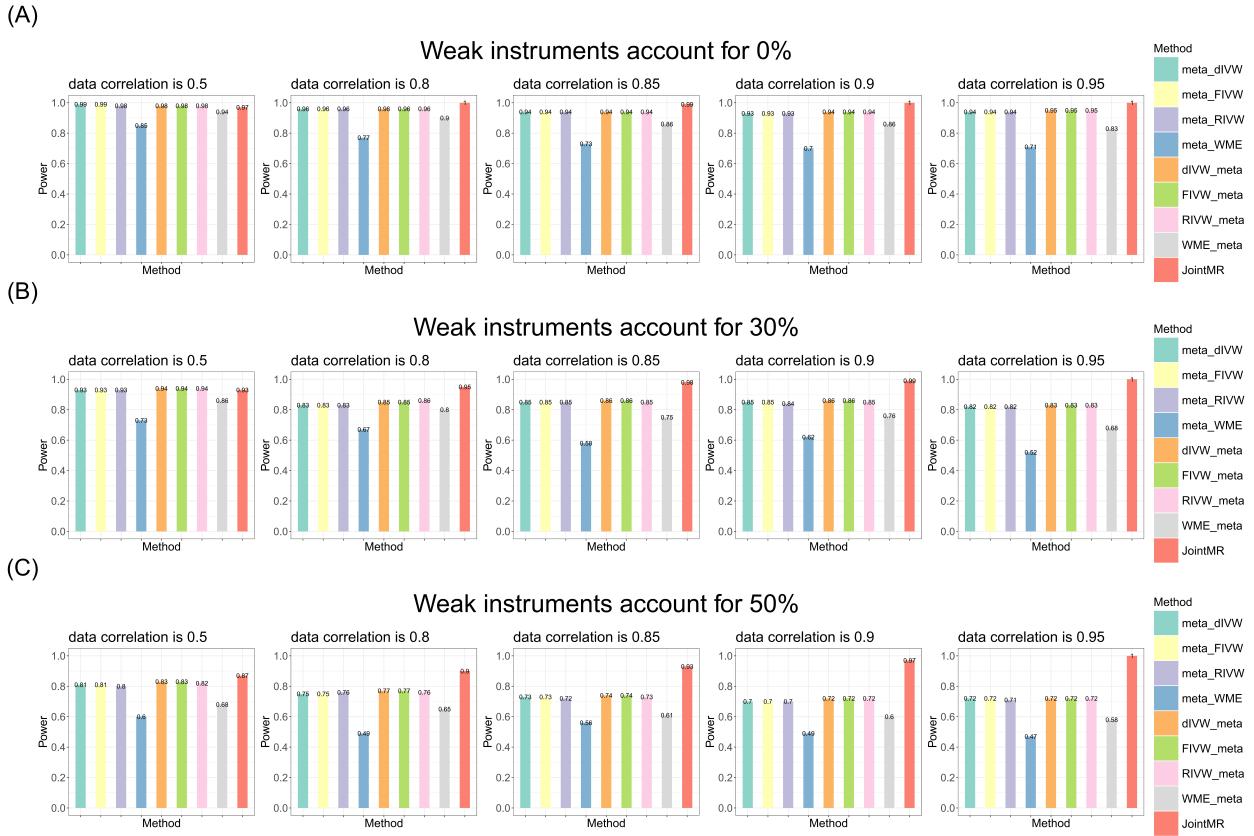
**Figure S6. Simulation results for statistical power by proportion of weak instruments (causal effect = 0.10).**

Bar charts show the statistical power for detecting a non-zero causal effect (true effect = 0.10) under scenarios with varying proportions of weak instruments: (A) 0%, (B) 30%, and (C) 50%.

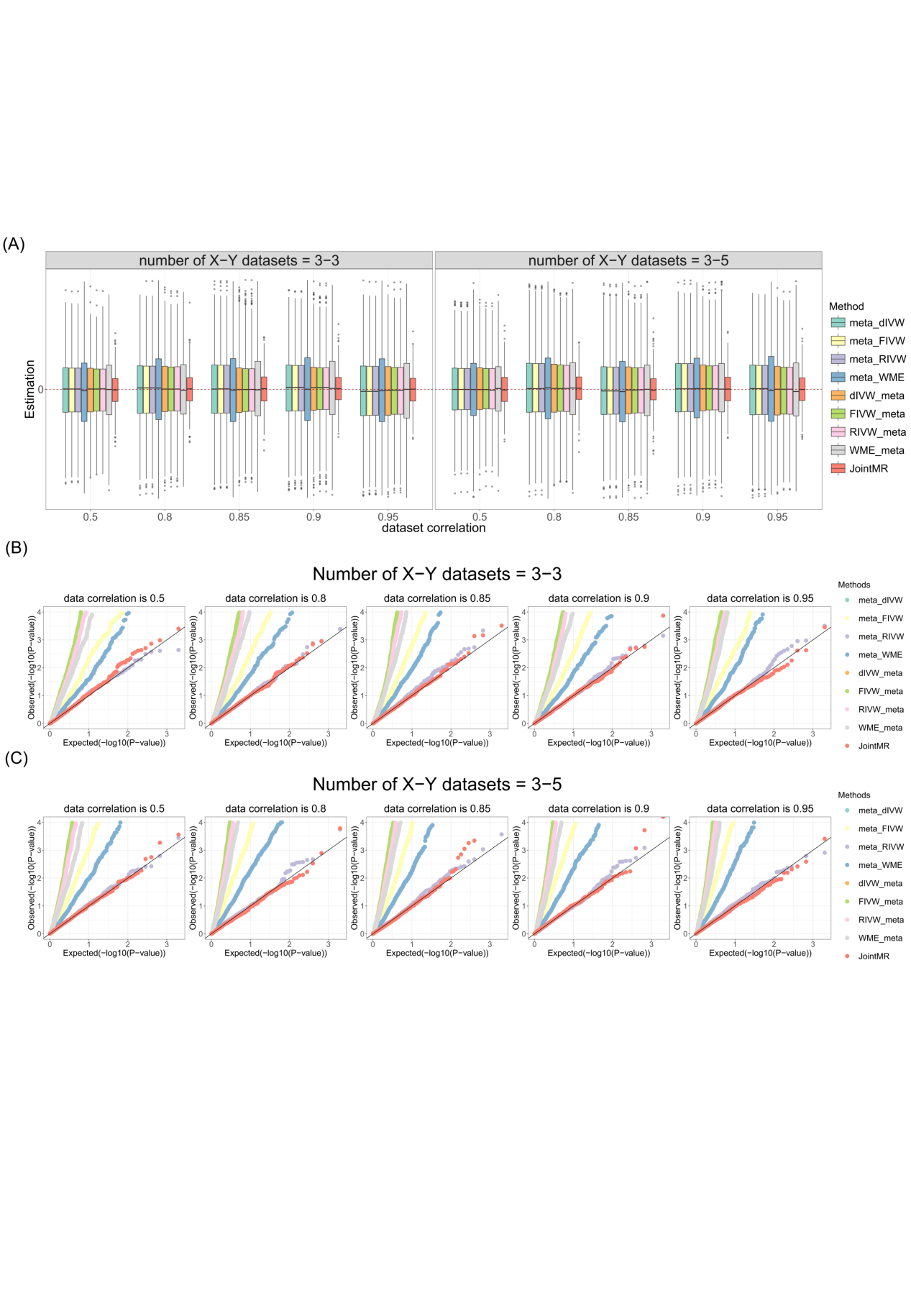

**Figure S7. Simulation results for causal effect estimation under the null scenario with random-effect model (no pleiotropy).**

(A) Boxplots show the performances of causal effect estimation when the true causal effect is zero. (B) and (C) Q-Q plots show the performances of Type I error rates for testing the null causal effect when the number of G-X and G-Y databases is 3 and 3 (B) and 3 and 5 (C), respectively.

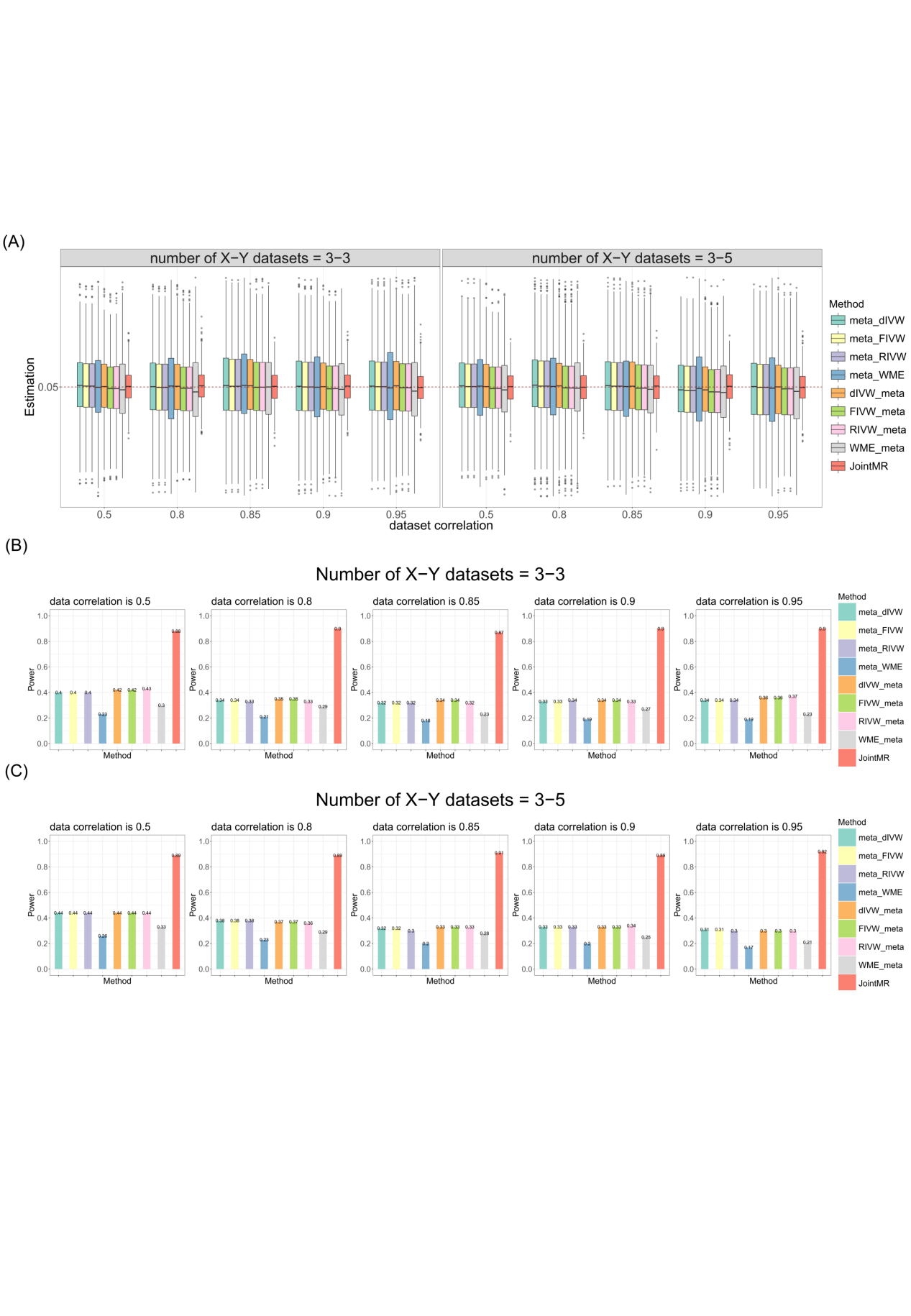

**Figure S8. Simulation results for causal effect estimation and statistical power under a non-null scenario with random-effect model (causal effect = 0.05, no pleiotropy).**

(A) Boxplots show the performances of causal effect estimation when the true causal effect is 0.05. (B) and (C) Bar charts show the statistical power for detecting the non-zero causal effect when the number of G-X and G-Y databases is 3 and 3 (B) and 3 and 5 (C), respectively.

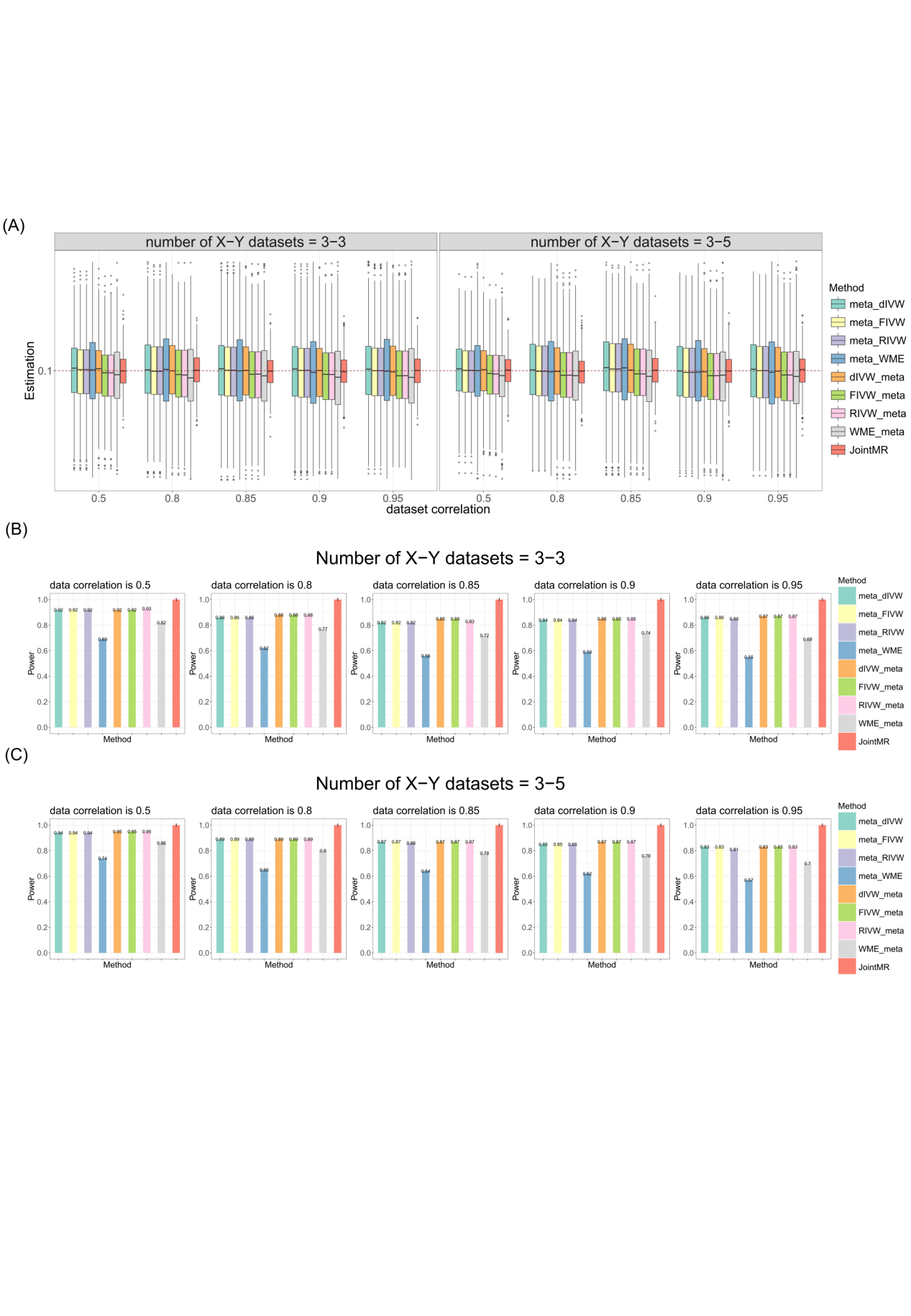

**Figure S9. Simulation results for causal effect estimation and statistical power under a non-null scenario with random-effect model (causal effect = 0.10, no pleiotropy).**

(A) Boxplots show the performances of causal effect estimation when the true causal effect is 0.1. (B) and (C) Bar charts show the statistical power for detecting the non-zero causal effect when the number of G-X and G-Y databases is 3 and 3 (B) and 3 and 5 (C), respectively.

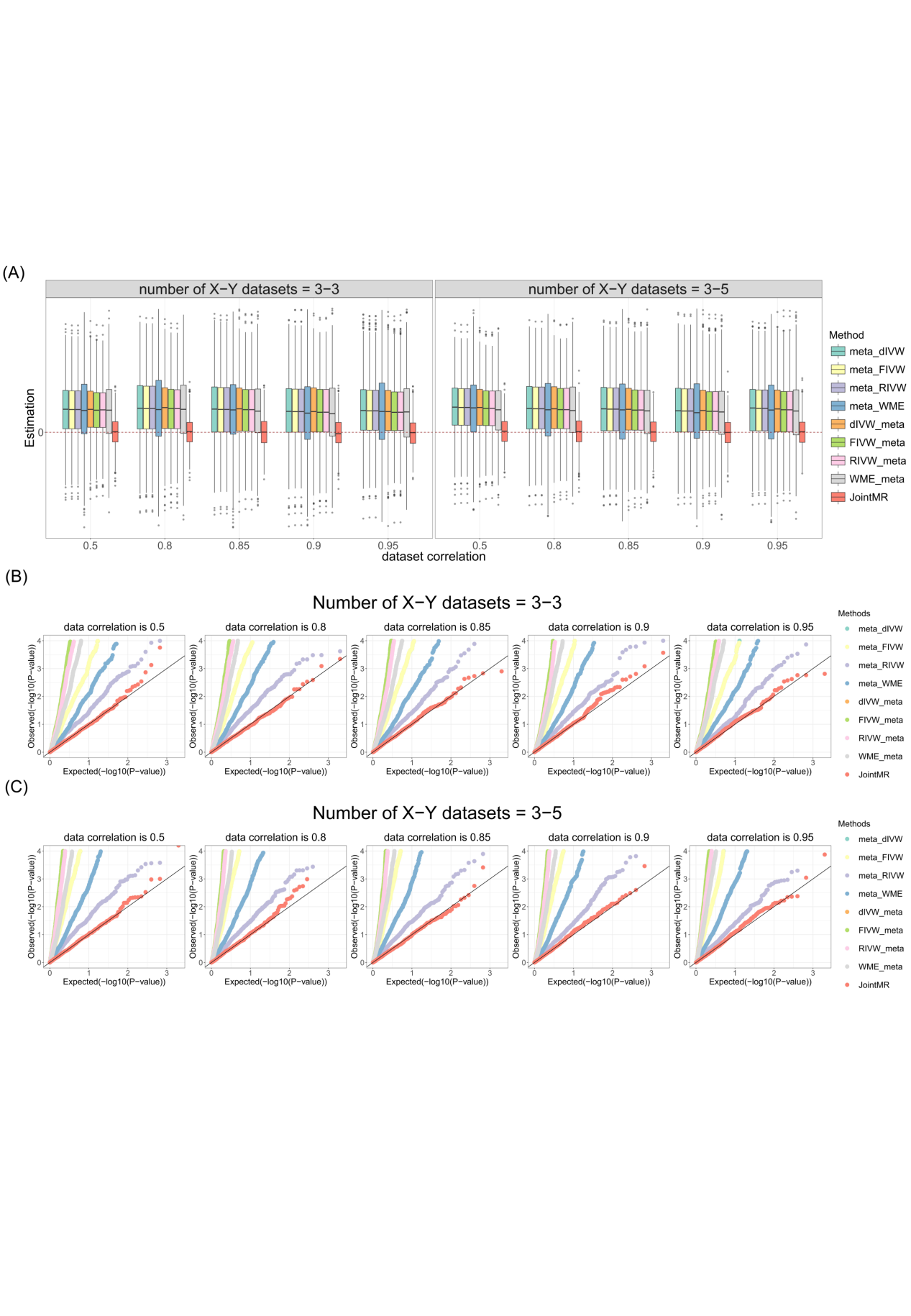

**Figure S10. Simulation results for causal effect estimation under the null scenario with random-effect model (horizontal pleiotropy).**

(A) Boxplots show the performances of causal effect estimation when the true causal effect is zero. (B) and (C) Q-Q plots show the performances of Type I error rates for testing the null causal effect when the number of G-X and G-Y databases is 3 and 3 (B) and 3 and 5 (C), respectively.

**Figure S11. Simulation results for causal effect estimation and statistical power under a non-null scenario with random-effect model (causal effect = 0.05, horizontal pleiotropy).**

(A) Boxplots show the performances of causal effect estimation when the true causal effect is 0.05. (B) and (C) Bar charts show the statistical power for detecting the non-zero causal effect when the number of G-X and G-Y databases is 3 and 3 (B) and 3 and 5 (C), respectively.

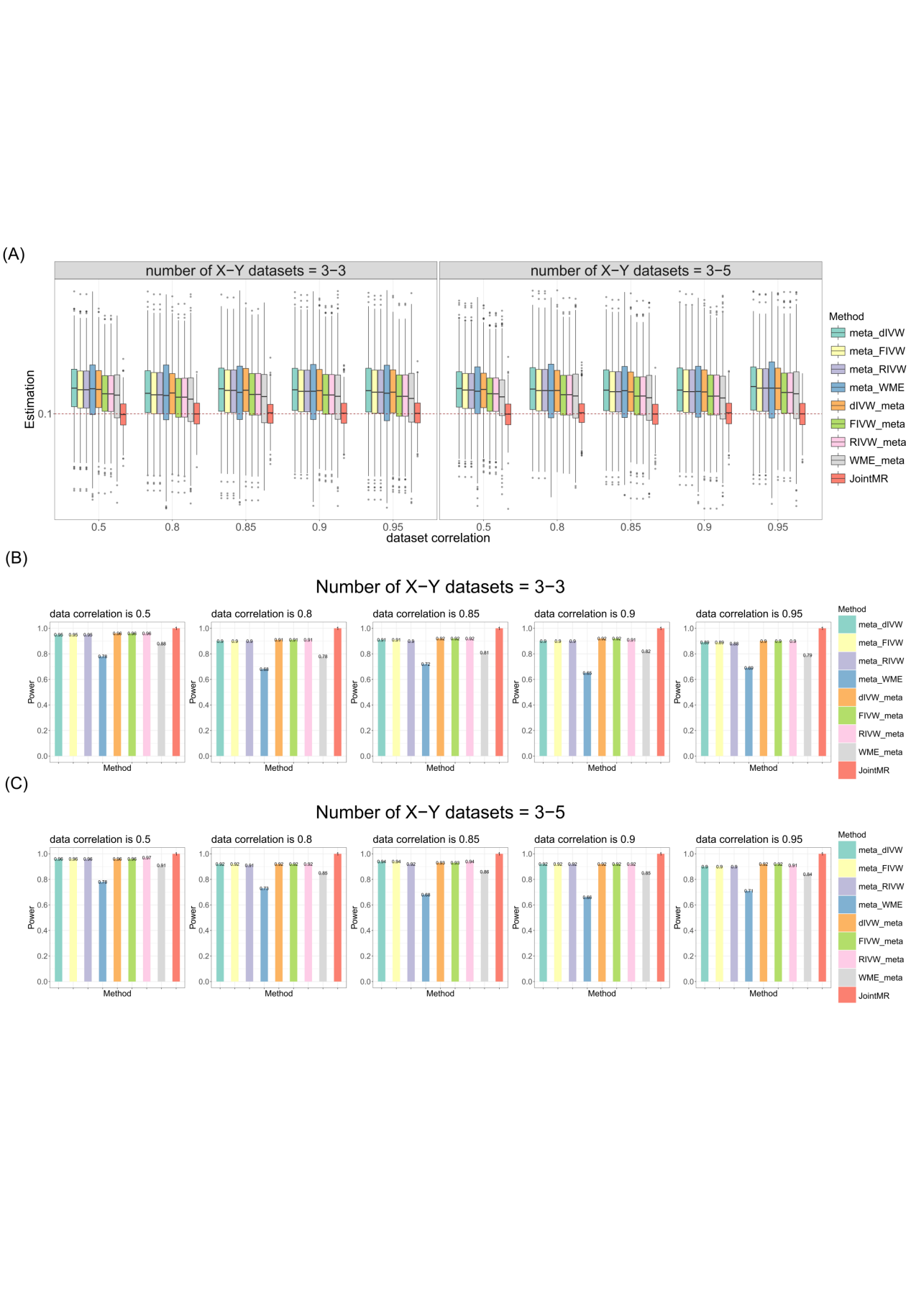

**Figure S12. Simulation results for causal effect estimation and statistical power under a non-null scenario with random-effect model (causal effect = 0.10, horizontal pleiotropy).**

(A) Boxplots show the performances of causal effect estimation when the true causal effect is 0.1. (B) and (C) Bar charts show the statistical power for detecting the non-zero causal effect when the number of G-X and G-Y databases is 3 and 3 (B) and 3 and 5 (C), respectively.

**Table S1 Information of application datasets**

| **Phenotype** | **Author** | **Id** | **Publication Date** | **Journal** | **Sample** | **Pubmed Id** | **Database** | **Dowanload site** |
| --- | --- | --- | --- | --- | --- | --- | --- | --- |
| T2D | Kurki MI | / | 2023/1/18 | Nature | 486367 | 36653562 | Finn | https://storage.googleapis.com/finngen-public-data-r12/summary_stats/release/finngen_R12_T2D.gz |
|  | Cai L | GCST90006934 | 2020/11/13 | Sci Data | 22326 | 33188205 | EPIC-InterAct project | http://ftp.ebi.ac.uk/pub/databases/gwas/summary_statistics/GCST90006001-GCST90007000/GCST90006934 |
|  | Mansour Aly D | GCST90026417 | 2021/11/4 | Nat Genet | 12230 | 34737425 | Swedish ANDIS | http://ftp.ebi.ac.uk/pub/databases/gwas/summary_statistics/GCST90026001-GCST90027000/GCST90026417 |
|  | Verma A | GCST90475667 | 2024/7/19 | Science | 432648 | 39024449 | MVP | http://ftp.ebi.ac.uk/pub/databases/gwas/summary_statistics/GCST90475001-GCST90476000/GCST90475667 |
|  | Loh PR | GCST90029024 | 2018/7/1 | Nat Genet | 468298 | 29892013 | UKB | <http://ftp.ebi.ac.uk/pub/databases/gwas/summary_statistics/GCST90029001-GCST90030000/GCST90029024> |
| TC | Willer CJ | GCST002221 | 2013/10/6 | Nat Genet | 93982 | 24097068 | GLGC | [http://ftp.ebi.ac.uk/pub/databases/gwas/summary_statistics/GCST002001-GCST003000/GCST002221/jointGwasMc_TC.txt.gz](http://ftp.ebi.ac.uk/pub/databases/gwas/summary_statistics/GCST002001-GCST003000/GCST002221/jointGwasMc_TC.txt.gz" \o "http://ftp.ebi.ac.uk/pub/databases/gwas/summary_statistics/GCST002001-GCST003000/GCST002221/jointGwasMc_TC.txt.gz) |
|  | Barton AR | GCST90025953 | 2021/7/5 | Nat Genet | 437878 | 34226706 | UKB | [http://ftp.ebi.ac.uk/pub/databases/gwas/summary_statistics/GCST90025001-GCST90026000/GCST90025953/GCST90025953_buildGRCh37.tsv](http://ftp.ebi.ac.uk/pub/databases/gwas/summary_statistics/GCST90025001-GCST90026000/GCST90025953/GCST90025953_buildGRCh37.tsv" \o "http://ftp.ebi.ac.uk/pub/databases/gwas/summary_statistics/GCST90025001-GCST90026000/GCST90025953/GCST90025953_buildGRCh37.tsv) |
| TG | Willer CJ | GCST002216 | 2013/10/6 | Nat Genet | 94595 | 24097068 | GLGC | <http://ftp.ebi.ac.uk/pub/databases/gwas/summary_statistics/GCST002001-GCST003000/GCST002216/jointGwasMc_TG.txt.gz> |
|  | Barton AR | GCST90025957 | 2021/7/5 | Nat Genet | 437532 | 34226706 | UKB | [http://ftp.ebi.ac.uk/pub/databases/gwas/summary_statistics/GCST90025001-GCST90026000/GCST90025957/GCST90025957_buildGRCh37.tsv](http://ftp.ebi.ac.uk/pub/databases/gwas/summary_statistics/GCST90025001-GCST90026000/GCST90025957/GCST90025957_buildGRCh37.tsv" \o "http://ftp.ebi.ac.uk/pub/databases/gwas/summary_statistics/GCST90025001-GCST90026000/GCST90025957/GCST90025957_buildGRCh37.tsv) |
| HDL-C | Barton AR | GCST90025956 | 2021/7/5 | Nat Genet | 400754 | 34226706 | UKB | <http://ftp.ebi.ac.uk/pub/databases/gwas/summary_statistics/GCST90025001-GCST90026000/GCST90025956/GCST90025956_buildGRCh37.tsv> |
|  | Verma A | GCST90475352 | 2024/7/19 | Science | 404121 | 39024449 | MVP | http://ftp.ebi.ac.uk/pub/databases/gwas/summary_statistics/GCST90475001-GCST90476000/GCST90475352 |
| LDL-C | Willer CJ | GCST002222 | 2013/10/6 | Nat Genet | 93982 | 24097068 | GLGC | http://ftp.ebi.ac.uk/pub/databases/gwas/summary_statistics/GCST002001-GCST003000/GCST002222 |
|  | Klimentidis YC | GCST90002412 | 2020/10/1 | Diabetes | 431167 | 32493714 | UKB | <http://ftp.ebi.ac.uk/pub/databases/gwas/summary_statistics/GCST90002001-GCST90003000/GCST90002412/GCST90002412_buildGRCh37.tsv.gz> |

**Table S2 Results of application**

| Exposure | Study | OR | SE(OR) | P-value | Type | n(SNP) |
| --- | --- | --- | --- | --- | --- | --- |
| TC | Single MR UKB-UKB | 0.986 | 0.002 | 5.409E-17 | Individual | 732 |
|  | Single MR UKB-FinnGen | 0.890 | 0.028 | 1.839E-04 | Individual | 728 |
|  | Single MR UKB-MVP | 0.843 | 0.029 | 9.503E-07 | Individual | 715 |
|  | Single MR UKB-EPIC | 0.842 | 0.048 | 2.674E-03 | Individual | 727 |
|  | Single MR UKB-Swedish | 0.796 | 0.068 | 7.118E-03 | Individual | 684 |
|  | Single MR GLGC-UKB | 0.995 | 0.002 | 5.169E-03 | Individual | 90 |
|  | Single MR GLGC-FinnGen | 0.966 | 0.033 | 3.110E-01 | Individual | 90 |
|  | Single MR GLGC-MVP | 0.914 | 0.032 | 1.072E-02 | Individual | 86 |
|  | Single MR GLGC-EPIC | 0.874 | 0.05 | 1.937E-02 | Individual | 88 |
|  | Single MR GLGC-Swedish | 0.859 | 0.086 | 1.286E-01 | Individual | 86 |
|  | GWAS-meta MR | 1.006 | 0.002 | 1.368E-02 | Summary | 206 |
|  | Traditional MR-meta | 0.911 | 0.022 | 1.425E-04 | Summary | - |
|  | JointMR | 1.236 | 0.029 | 8.212E-20 | Summary | 207 |
| TG | Single MR UKB-UKB | 1.014 | 0.002 | 1.545E-16 | Individual | 1125 |
|  | Single MR UKB-FinnGen | 1.304 | 0.044 | 3.819E-15 | Individual | 1121 |
|  | Single MR UKB-MVP | 1.419 | 0.046 | 1.623E-27 | Individual | 1106 |
|  | Single MR UKB-EPIC | 1.385 | 0.073 | 6.875E-10 | Individual | 1121 |
|  | Single MR UKB-Swedish | 1.533 | 0.120 | 5.348E-08 | Individual | 1065 |
|  | Single MR GLGC-UKB | 1.002 | 0.004 | 6.176E-01 | Individual | 53 |
|  | Single MR GLGC-FinnGen | 1.051 | 0.086 | 5.446E-01 | Individual | 53 |
|  | Single MR GLGC-MVP | 1.057 | 0.082 | 4.755E-01 | Individual | 52 |
|  | Single MR GLGC-EPIC | 1.164 | 0.113 | 1.178E-01 | Individual | 53 |
|  | Single MR GLGC-Swedish | 1.105 | 0.157 | 4.807E-01 | Individual | 50 |
|  | GWAS-meta MR | 0.991 | 0.003 | 1.845E-03 | Summary | 305 |
|  | Traditional MR-meta | 1.192 | 0.062 | 6.893E-04 | Summary | - |
|  | JointMR | 1.306 | 0.060 | 6.607E-09 | Summary | 1000 |
| HDL-C | Single MR UKB-UKB | 0.986 | 0.001 | 5.154E-32 | Individual | 1616 |
|  | Single MR UKB-FinnGen | 0.812 | 0.017 | 3.138E-22 | Individual | 1625 |
|  | Single MR UKB-MVP | 0.763 | 0.016 | 6.468E-36 | Individual | 1628 |
|  | Single MR UKB-EPIC | 0.759 | 0.030 | 4.761E-12 | Individual | 1613 |
|  | Single MR UKB-Swedish | 0.717 | 0.048 | 5.533E-07 | Individual | 1317 |
|  | Single MR MVP-UKB | 0.990 | 0.001 | 4.926E-52 | Individual | 1118 |
|  | Single MR MVP-FinnGen | 0.831 | 0.011 | 8.483E-44 | Individual | 1117 |
|  | Single MR MVP-MVP | 0.728 | 0.010 | 4.873E-120 | Individual | 1119 |
|  | Single MR MVP-EPIC | 0.793 | 0.018 | 4.029E-24 | Individual | 1113 |
|  | Single MR MVP-Swedish | 0.771 | 0.029 | 6.735E-12 | Individual | 938 |
|  | GWAS-meta MR | 1.007 | 0.002 | 4.620E-04 | Summary | 439 |
|  | Traditional MR-meta | 0.814 | 0.030 | 2.276E-08 | Summary | - |
|  | JointMR | 0.904 | 0.024 | 1.536E-04 | Summary | 3004 |
| LDL-C | Single MR UKB-UKB | 1.000 | 0.002 | 9.894E-01 | Individual | 420 |
|  | Single MR UKB-FinnGen | 1.056 | 0.030 | 5.864E-02 | Individual | 417 |
|  | Single MR UKB-MVP | 1.062 | 0.035 | 6.735E-02 | Individual | 416 |
|  | Single MR UKB-EPIC | 0.991 | 0.048 | 8.551E-01 | Individual | 417 |
|  | Single MR UKB-Swedish | 0.910 | 0.073 | 2.390E-01 | Individual | 395 |
|  | Single MR GLGC-UKB | 1.000 | 0.001 | 7.685E-01 | Individual | 436 |
|  | Single MR GLGC-FinnGen | 1.050 | 0.023 | 2.603E-02 | Individual | 430 |
|  | Single MR GLGC-MVP | 1.053 | 0.025 | 2.886E-02 | Individual | 428 |
|  | Single MR GLGC-EPIC | 1.003 | 0.040 | 9.498E-01 | Individual | 435 |
|  | Single MR GLGC-Swedish | 0.936 | 0.066 | 3.470E-01 | Individual | 399 |
|  | GWAS-meta MR | 1.001 | 0.002 | 7.143E-01 | Summary | 441 |
|  | Traditional MR-meta | 1.017 | 0.011 | 9.951E-02 | Summary | - |
|  | JointMR | 1.302 | 0.045 | 1.509E-14 | Summary | 1552 |
